# Use of HIV Recency Assays for HIV Incidence Estimation and Non-Incidence Surveillance Use Cases: A systematic review

**DOI:** 10.1101/2021.08.23.21262504

**Authors:** Shelley N. Facente, Lillian Agyei, Andrew D. Maher, Mary Mahy, Shona Dalal, David Lowrance, Eduard Grebe, Kimberly Marsh

## Abstract

**Introduction:** HIV assays designed to detect recent infection, also known as “recency assays,” are often used to estimate HIV incidence in a specific country, region, or subpopulation, alone or as part of recent infection testing algorithms (RITAs). Recently, many countries and organizations have become interested in using recency assays within case surveillance systems and routine HIV testing services, and in measuring other indicators beyond incidence, generally referred to as “non-incidence surveillance use cases.”

**Methods:** To identify best methodological and field implementation practices for the use of recency assays to estimate HIV incidence and trends in recent infections for key populations or specific geographic areas, we undertook: 1) a global Call for Information released from WHO/UNAIDS; and 2) a systematic review of the literature to: (a) assess the field performance characteristics of commercially available recency assays, (b) understand the use of recency testing for surveillance in programmatic and laboratory settings, and (c) review methodologies for implementing recency testing for both incidence estimation and non-incidence use cases.

**Results and discussion:** Among the 90 documents ultimately reviewed, 65 (88%) focused on assay/algorithm performance or methodological descriptions, with high-quality evidence of accurate age- and sex- disaggregated HIV incidence estimation at national or regional levels in general population settings, but not at finer geographic levels for prevention prioritization. The remaining 25 documents described field-derived incidence (n=14) and non-incidence (n=11) use cases, including integrating RITAs into routine surveillance and assisting with molecular genetic analyses, but evidence was generally weaker or only reported on what was done, without validation data or findings related to effectiveness of recency assays when used for these purposes.

**Conclusions:** HIV recency assays have been widely validated for estimating HIV incidence in age- and sex-specific populations at national and sub-national regional levels; however, there was a lack of evidence validating the accuracy and effectiveness of using recency assays for non-incidence surveillance use cases. The evidence identified through this review will be used in forthcoming technical guidance on the use of HIV recency assays for surveillance use cases by WHO and UNAIDS; further evidence on methodologies and effectiveness of non-incidence use cases is needed.

## Introduction

There are many reasons to identify recently acquired HIV infections on a population level, including: (1) To better understand current transmission of HIV in a country, region, or population subgroup, (2) To evaluate whether specific prevention interventions are having the desired impact, and (3) To focus limited resources for prevention or treatment services on groups of people or geographic locations with the greatest potential benefit (e.g., reducing risk for onward transmission). HIV assays designed to detect recent infection, also known as “recency assays,” can be used to gain an understanding of these epidemic dynamics.

Recency assays discriminate recent from longstanding infection in an individual using one or more biomarkers, typically using an understanding of the typical patterns of immune response maturation following initial infection. Individual recency assay results can be used in a cross-sectional survey to estimate incidence by building on the common epidemiological equation *P = I × D* (prevalence = incidence × duration of infection).^1^ However, the accuracy of the incidence estimate is dependent upon accurate knowledge of the performance characteristics of the recency assay or algorithm, specifically mean duration of recent infection (the average time post-infection that individuals are classified as recently infection; MDRI) and false-recent rate (the proportion of long-infected individuals misclassified as recently infected; FRR), and the precision of the estimate is sensitive to these same parameters.^2^

To date, no recency assay has fully met the target product profile for HIV incidence estimation as set out by the Foundation for Innovative Diagnostics (FIND) and the World Health Organisation (WHO) in 2016.^3,4^ Numerous factors have been identified that adversely affect recency assay performance and lead to substantial misclassification of longstanding infections as recent (i.e., raise the FRR). Factors that can affect assay performance include natural variability in individual immune responses (in particular, elite control of HIV or natural viral suppression), variability in biomarker progression for different HIV-1 subtypes, the types of specimens collected and storage methods, advanced HIV disease, and treatment with antiretroviral therapy (ART) or use of pre-exposure prophylaxis (PrEP).^5-10^ The effect of ART on increasing the FRR of recency assays appears to be more pronounced when a person receives treatment very early after initial infection;^11,12^ which is complicated by rapid improvements in treatment coverage worldwide, as well as uptake of pre-exposure prophylaxis (PrEP). Other factors that may impact assay performance but are not yet well-characterised include sex, pregnancy status and the presence of co-morbidities.^13-15^

Since the release in 2011 of technical guidance on the use of recency assays to estimate population-level HIV incidence from the WHO and Joint United Nations Programme on HIV/AIDS (UNAIDS),^16^ the field has changed substantially, motivating release of interim guidance at various times.^13,17-20^ Numerous examples in the peer-reviewed literature now highlight the necessity of adjustments at a local level to improve the accuracy of incidence estimates derived using recency assays within population-based surveys.^14,21-30^ Beyond that primary application, however, many countries and organizations have become increasingly interested in using recency assays within HIV case surveillance systems and routine HIV testing services, to measure indicators other than incidence, such as the identification of epidemiologically-linked clusters of recent infections; geographic hotspots; or subpopulations with relatively high, ongoing, or emerging transmission, to inform prioritization of HIV prevention, testing, and partner notification or contact tracing interventions. These types of epidemic monitoring and evaluation strategies are generally referred to as “non-incidence surveillance use cases” for recency assays. However, the non-random nature by which people are included in these types of surveillance systems and programmes requires special attention to characterize and, ideally, mitigate the effect of these selection biases on the accuracy of these non-incidence estimates.

## METHODS

To identify methodological and field implementation practices for the use of recency assays for HIV incidence and non-incidence surveillance use cases, we used two strategies. First, a global Call for Information was released as a joint endeavour between WHO and UNAIDS staff working on both surveillance and HIV testing services. This call included a brief questionnaire through which member states and/or regional health jurisdictions were asked to describe current implementation (if any) of recency testing in programmatic HIV testing services in terms of assay performance, clinical utility, and utility for surveillance, including but not limited to incidence estimation.

Second, a systematic review of the literature was conducted, with four primary objectives:

1. Understand the use of recency testing in surveillance, programmatic and laboratory settings (to provide incidence estimates or for non-incidence surveillance use cases),
2. Review methodologies for implementing recency testing in population surveys, case surveillance systems and routine monitoring & evaluation activities, and
3. Highlight use cases that have employed a recency assay or Recent Infection Testing Algorithm (RITA) within specific populations, with special attention to variations in assays, settings, and methods of analysis for calculating HIV incidence estimates and/or employing recency assays for non-incidence surveillance use cases.

### Eligibility Criteria for the Systematic Review

The systematic review included two sets of searches, each with a different strategy. Strategy 1 involved looking for articles about recency assay performance in laboratory and field survey settings. To be eligible for inclusion in the review, articles needed to describe some aspect of performance of recency assays/methodologies (e.g., MDRI, FRR, sensitivity, specificity, false positive rate, false negative rate, accuracy, number tested and proportion recently infected; or correlation, R, percent agreement, or kappa related to another standard assay). They also needed to use commercially available assays/methodologies used to determine recency of infection (see **Table 1** for this list). Articles reviewing use of a laboratory-developed (“home-grown”) assay that was not commercially available were excluded from the review.

**Table 1.** List of commercially available recency assays at the time of the review.

| Product name (manufacturer) | Assay type |
| --- | --- |
| Asante™ HIV-1 Rapid Recency® Assay (Sedia Biosciences) | Rapid, point-of-care |
| HIV Swift Recent Infection Assay (Maxim Biomedical) | Rapid, point-of-care |
| Sedia™ HIV-1 Limiting Antigen Avidity (LAg-Avidity) EIA (Sedia Biosciences) | Laboratory-based |
| Maxim HIV-1 LAg-Avidity EIA Kit (Maxim Biomedical) | Laboratory-based |
| Genetics Systems HIV-1/HIV-2 Plus O EIA (Bio-Rad, avidity protocol) | Laboratory-based |
| ARCHITECT HIV Ag/Ab Combo (Abbott, avidity protocol or unmodified protocol) | Laboratory-based |
| VITROS Anti-HIV 1+2 (Ortho Diagnostics, avidity protocol) | Laboratory-based |
| Geenius HIV -1/2 Confirmatory (Bio-Rad, modified protocol) | Laboratory-based |
| Inno-Lia® HIV I/II Score (Fujirebio, Inc.) | Laboratory-based |
| Sedia™ BED HIV-1 Incidence EIA (Sedia Biosciences) | Laboratory-based |

Strategy 2 involved looking for articles about surveillance and programmatic utilization of recency testing; articles reviewed under this strategy was intended to supplement findings from the WHO Call for Information as described above. To be eligible for inclusion, articles needed to describe some aspect of population-level utility (identification of “hotspots”, clusters, case surveillance and/or incidence estimation), using commercially available recency assays/methodologies (e.g., RITAs, adapted assay protocols) used to determine recency of HIV infection. Studies could present either qualitative or quantitative data, and could be descriptive studies lacking a comparator, as long as studies clearly presented outcomes specific to HIV recency testing.

### Search Strategy

The literature search for the systematic review was conducted in PubMed and Web of Science, and included literature published in any language and in any indexed journal including preprint servers without peer review, from 1 January 2010 to 5 January 2021, searching title, abstract, and MeSH Terms/author keywords.

For the Strategy 1 search, search terms included HIV; recency assay; incidence assay; test for recent infection (TRI); false recent rate/ratio (FRR); proportion false recent; and mean duration of recent infection (MDRI). For the Strategy 2 search, search terms included recent infection/acute infection; recent infection testing algorithm (RITA); incidence estimates; case surveillance; hotspot identification; hotspot mapping; cluster detection; procedures and protocols; and HIV. See Table S1 for search sets and terms, and Table S2 for our search code.

Given that much of the research output in the field of HIV recency assay utilization is published in formal reports or presented in conference abstracts, we extended the search beyond traditional literature databases to include “grey literature,” i.e., literature that is not formally published in peer-reviewed journals or books. We conducted a search of the grey literature through internet search engines and through websites of major international funders, subject matter conferences, and organizations involved with HIV surveillance (see Table S3) employing the following search terms across sites: “surveillance,” “recency testing,” “case surveillance,” “incidence estimation,” “hotspot,” and “HIV.”

We used a step-wise approach during the screening and reviewing process. After search and duplicate removal, LA screened titles and abstracts to identify papers potentially related to the focus areas and eligibility criteria. After screening was complete, remaining articles for full-text could be obtained were then independently reviewed by SNF and LA to determine if the study met eligibility criteria; ADM served as a tiebreaker for any articles for which the two preliminary screeners were not in agreement about inclusion. Once the full-text review was complete, LA hand-searched the references of all included articles for additional, potentially eligible articles. SNF and LA then reviewed these articles and determined eligibility according to the process outlined above.

Prior to conducting our search, we developed a formal protocol and circulated it among stakeholders at UNAIDS and WHO for approval; we have made the protocol available in unmodified form as supplemental material to this article.

### Assessment of Evidence Strength

Both protocols and reports submitted by countries as supporting documentation in response to the WHO Call for Information, and literature included in the systematic review was rated by strength of published evidence using a 23-point rubric (see **Figure 1**). For each piece of evidence, two team members (SNF and LA) independently rated the strength of evidence through a Microsoft Excel-based scoring rubric designed to implement the grading structure found in Figure 1. If there was disagreement between the two team members, either ADM or EG performed an assessment using the rubric and served as a tiebreaker.

**Figure 1.**
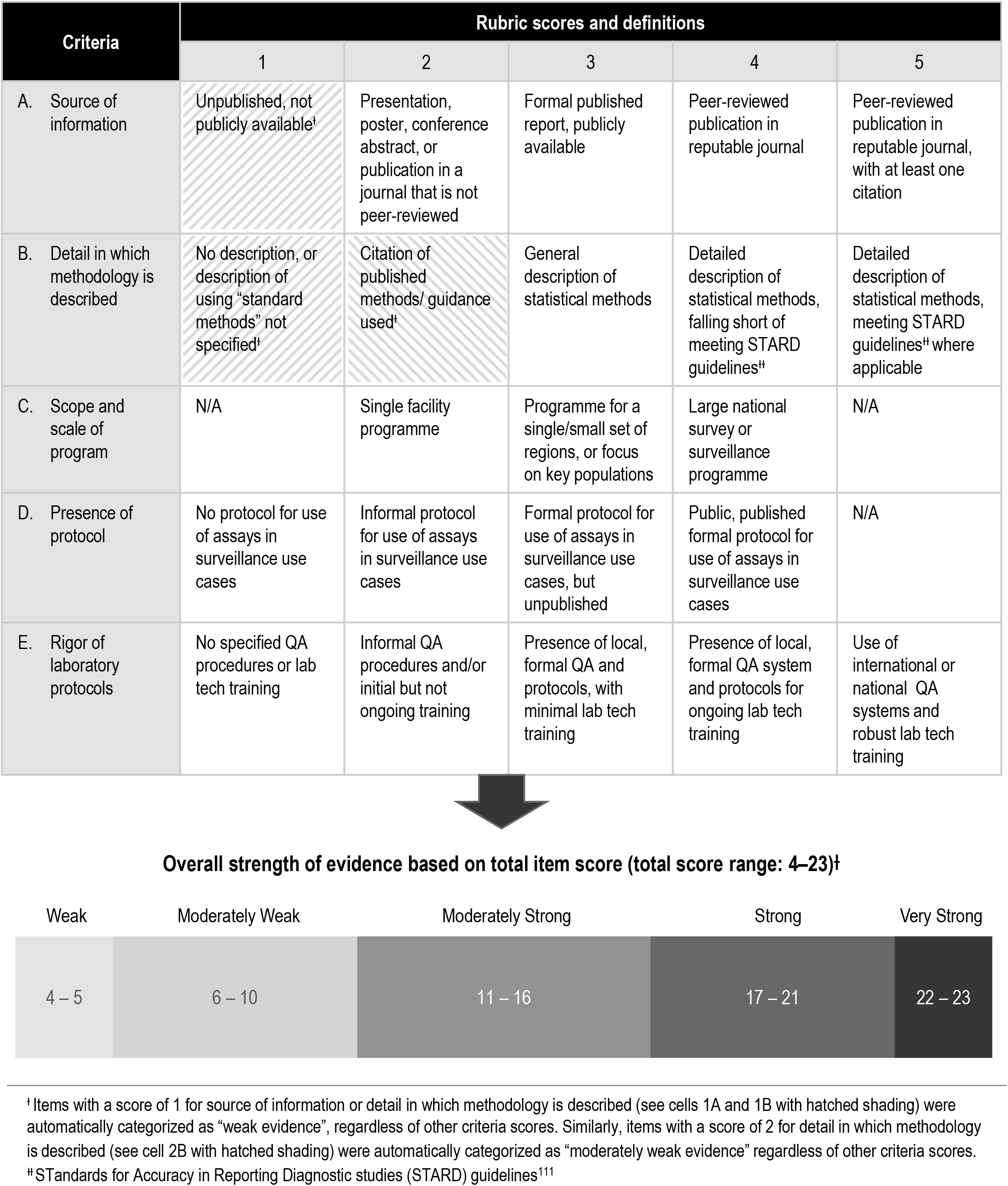
Rubric used to evaluate strength of evidence for each item reviewed. A score ranging from 1–5 was assigned to each item based on the following criteria: A) source of information, B) detail in which methodology was described, C) scope and scale of program, D) presence of protocol, and E) rigor of laboratory protocols. Each item was assigned an overall strength of evidence rating based on the sum of criteria scores^Ɨ^: weak evidence (overall score 4–5), moderately weak evidence (overall score 6–10), moderately strong evidence (overall score 11–16), strong evidence (overall score 17–21), and very strong evidence (overall score 22–23).

## RESULTS AND DISCUSSION

The search was conducted on 5 January 2021, and resulted in 611 records identified via MEDLINE (PubMed), 823 records identified via Web of Science, and 315 records identified through an Internet search of grey literature. An initial “quick screen” round resulted in 1,271 records from the MEDLINE and Web of Science searches being removed from the pool for clearly not meeting inclusion criteria for the review. Of the 478 pieces of evidence remaining after the quick screen, 58 records were identified as duplicate between the two databases and grey literature search and, once removed, resulted in 420 abstracts to be formally screened. Over a 4-month period ending 20 January 2021, 48 survey responses were received to the global Call for Information from five different WHO regions (AFRO, SEARO, WPRO, PAHO, and EURO).

### Literature screening steps

After de-duplication, a remaining 420 documents from the search were scanned by LA to identify papers related to our two strategy areas based on the eligibility criteria. A total of 380 records were then excluded; reasons for exclusion are detailed in **Figure 2**. The remaining 140 documents were then subjected to a full text review, which was conducted independently by both LA and SNF. After excluding full-text articles that did not meet our pre-defined inclusion criteria, a total of 74 studies, reports, or presentations were retained across both focus areas (**Figure 2**). An additional 16 pieces of evidence were then incorporated into the review from country submissions in response to the global Call for Information, leading to 90 items that were graded for strength of evidence.

**Figure 2.**
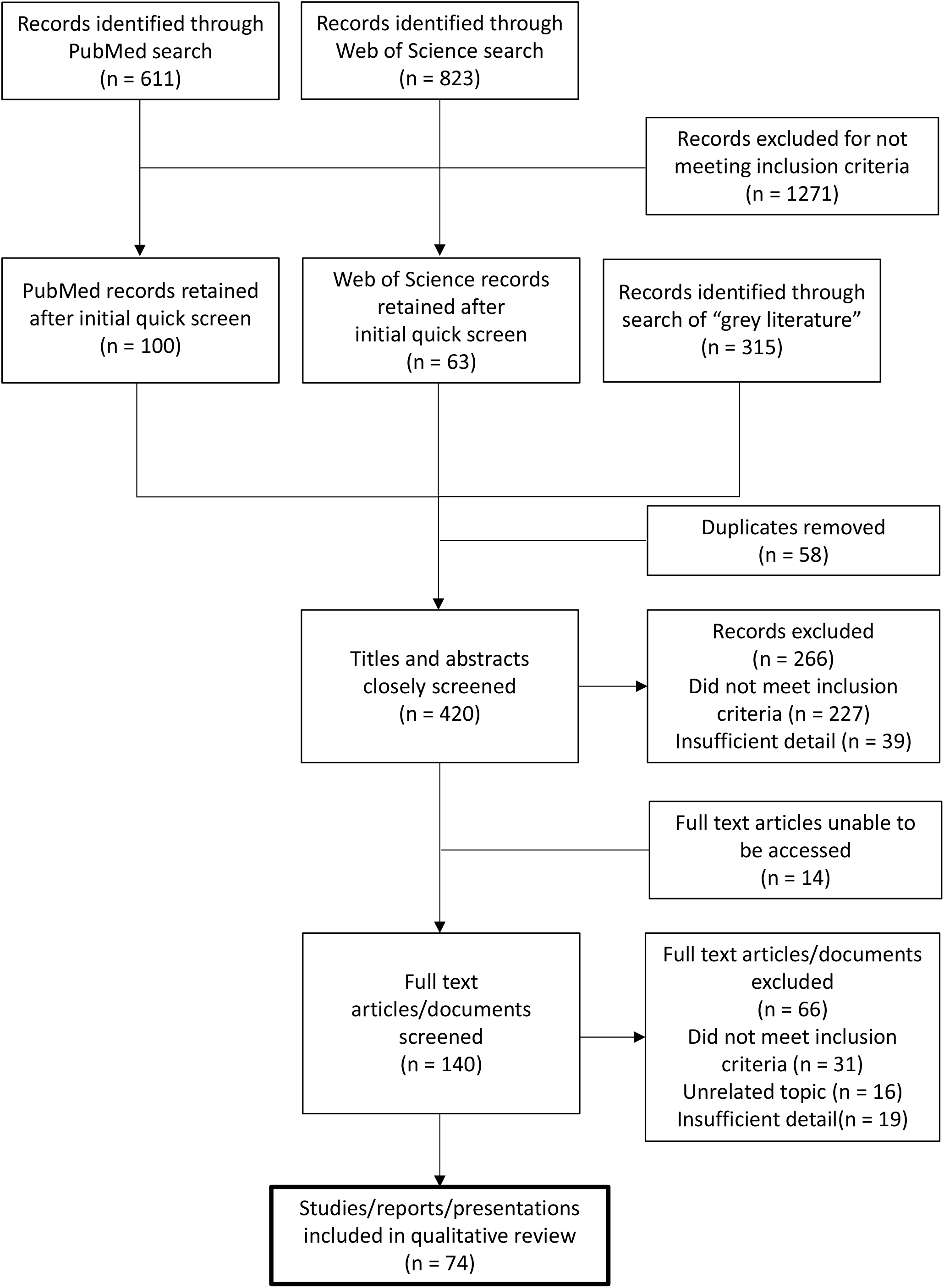
Flowchart of search process and results.

### Characteristics of Included Studies

Among the 90 pieces of evidence that were identified through country survey and systematic review and that met the inclusion criteria, 65 (88%) ^1,4,5,7-15,20,22-73^ focused on assay performance, algorithm performance, or methodological descriptions of incidence estimation. The quality of evidence was “very-strong” (40/65), “strong” (13/65), “moderately strong” (11/65), and “weak” (1/65) in these 65 articles. The remaining 25 pieces of evidence described field-derived incidence and non-incidence use cases. Of these, 16 (64%) described use for incidence estimation, 8 describe non-incidence use cases, and 1 described both incidence and non-incidence use cases.

Among the articles describing use of recency assays for estimation of HIV incidence, 12 (75%) ^74-85^ described *national surveillance* in the form of population-based surveys (including 10 from the US-supported Population-based HIV Impact Assessment (PHIA) surveys). These population-based incidence use cases are also sometimes known as *impact assessment* use cases, because they are intended for repeat implementation to assess changes in incidence over time as a result of HIV prevention or care interventions. Most evidence in this category (7/12) was judged to be “moderately strong,” with more details of strength ratings found in **Table 2**. The remaining five ^86-90^ articles described calculation of incidence among *key or sentinel populations*, including those accessing routine HIV or blood donation programs. Key or sentinel population surveillance involves testing within populations that are either of specific interest because they are at higher risk for infection (key) or considered to be representative of a larger population (sentinel). Sentinel and key population surveillance may be facility-based or community-based. For example, needle and syringe distribution programmes are a good point of contact with people who inject drugs, sexual health clinics may provide access to men who have sex with men (MSM) and sex workers, and antenatal clinics are used to sample pregnant women. Most evidence in this category (4/5) was “very strong” quality.

**Table 2.**
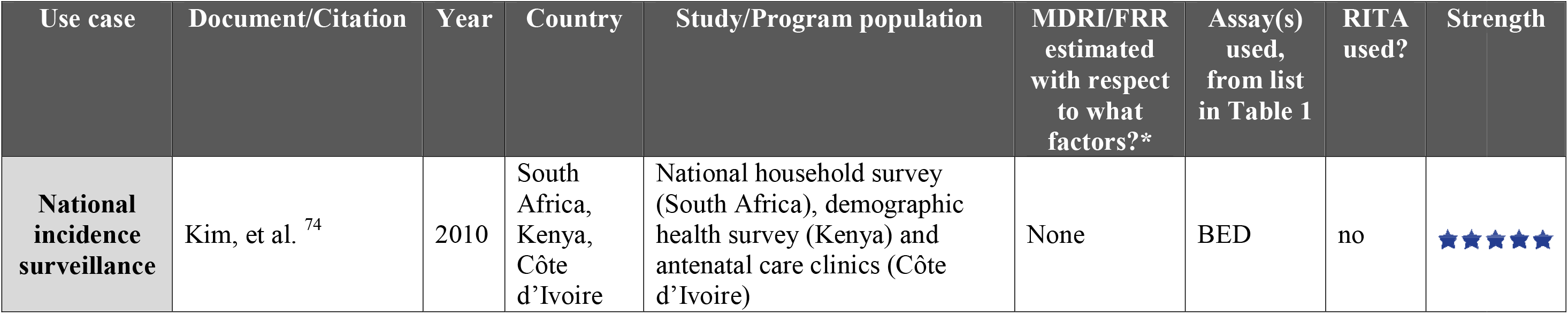
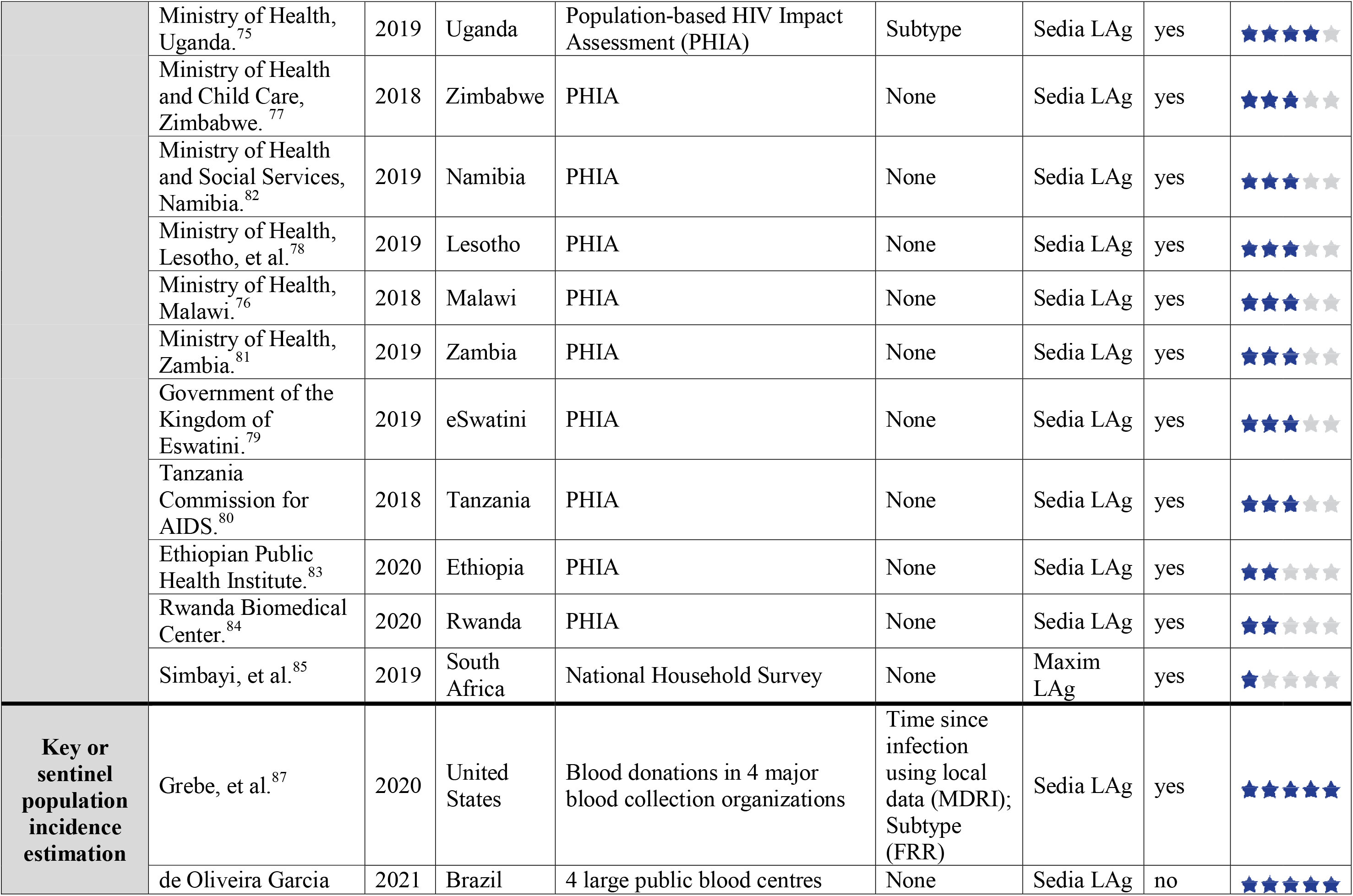
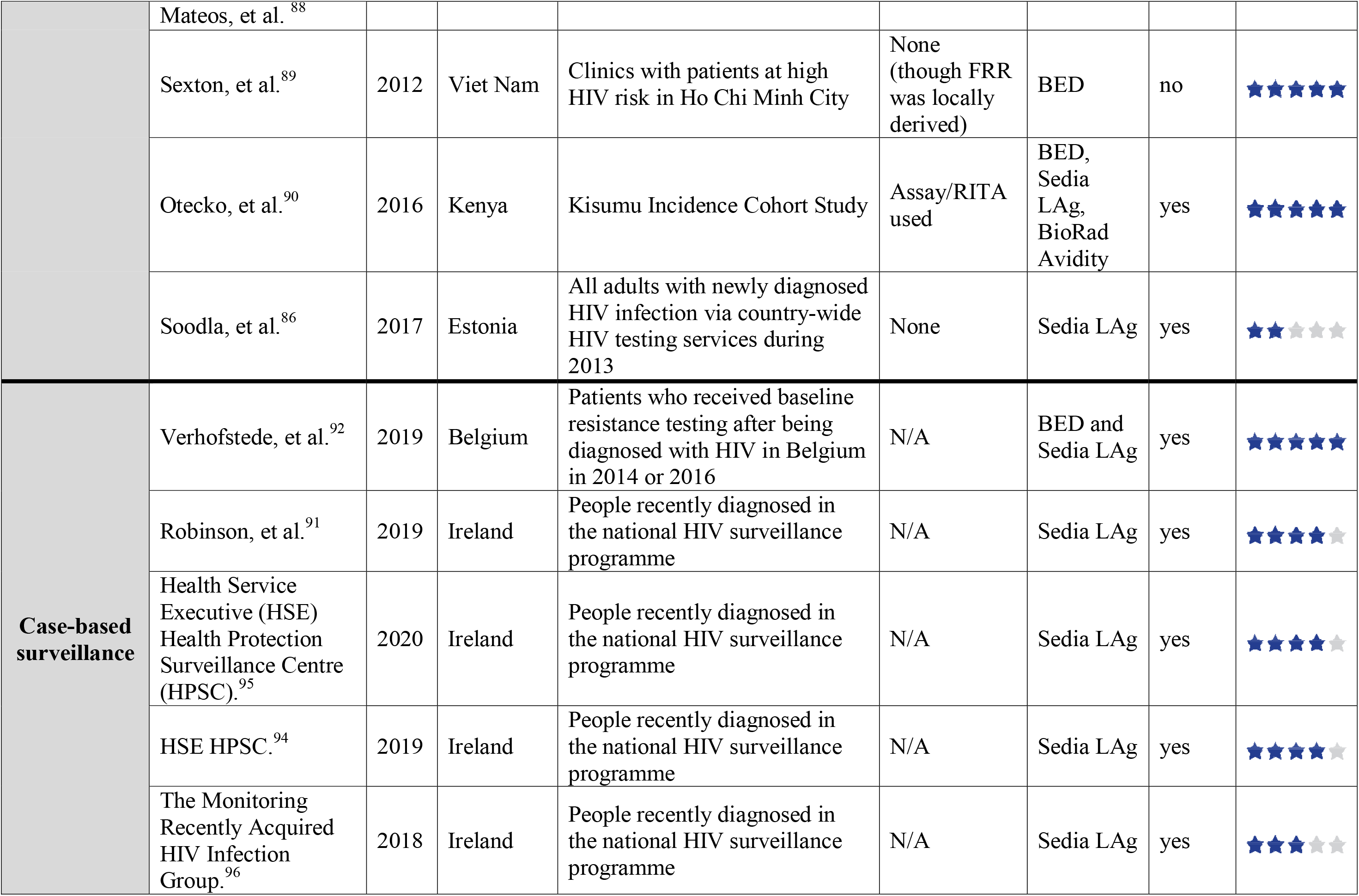
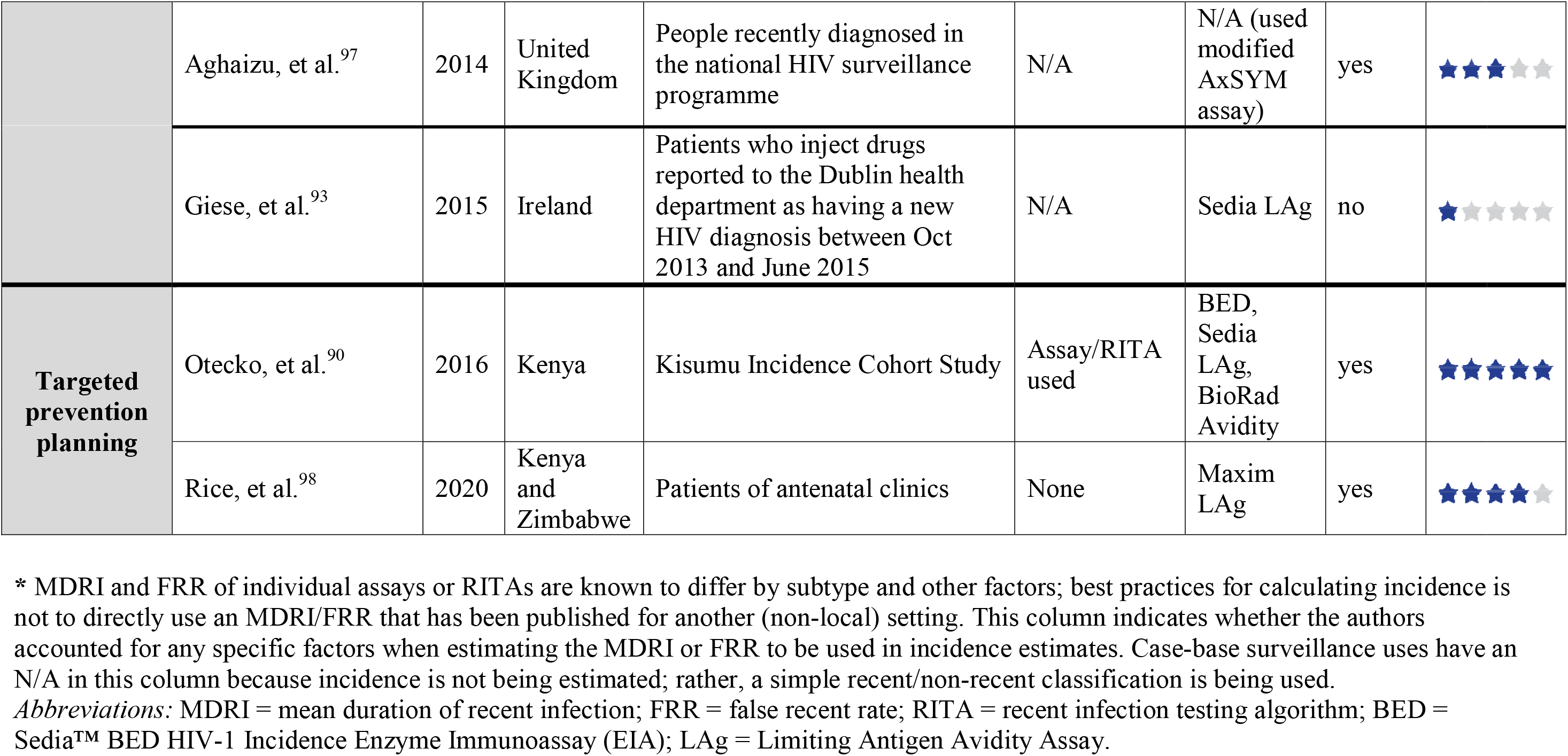
Summary of documents providing evidence related to use of recency assays under the four major surveillance use cases, from 1 January 2010 to 5 January 2021.

Among non-incidence use cases, seven^91-97^ described *case-based surveillance* among new HIV diagnoses in routine HIV programme settings and 2 described recency testing to inform *targeted prevention planning*, including one from a population-level cohort study^90^ and one that described use of recency testing within three separate routine HIV country programs.^98^ The quality of evidence was at least “moderately strong” in all 9/9 non-incidence use cases, each identified from the peer-reviewed literature. Each of these 9 articles used *proportion testing recent* and/or *odds for recently acquired infection* as the primary outcome measures.

Tables S4-S5 provide details on each of the 90 pieces of evidence included in this review, including the strength rating and topic of focus for each item.

### Use of Recency Assays for HIV Incidence Estimation

In 2015 UNAIDS and WHO released guidelines on monitoring the impact of the HIV epidemic using population-based surveys.^99^ Since then, 12 population-based surveys with published results have utilized this approach, the majority (n=10) of which were part of the global PHIA assessment.^100^ In addition to five articles that described efforts to estimate incidence within key or sentinel populations (including those accessing routine HIV programs^72,86,89^ or blood donation programs^87,88^), results from 10 completed PHIA surveys with public final reports were also entered into the review.^75-84^ These surveys involve cross-sectional, household-based, nationally representative sampling of adults and adolescents aged 15 years and older, with some surveys also including children aged 0-14 years. All PHIA countries were located in sub-Saharan Africa, except Haiti (which did not contribute evidence to this review).^101^ PHIA participants receive home-based HIV testing and counselling. Those who are HIV-positive undergo a laboratory-based RITA. During the first three PHIA surveys in Malawi, Zimbabwe, and Lesotho, the RITA included the Sedia™ HIV-1 Limiting Antigen (LAg) Avidity assay in combination with viral load (VL). The subsequent 7 surveys added antiretroviral (ARV) detection to the LAg and VL tests as an enhanced measure to distinguish recent from long-term infections. Incidence estimates were obtained from the RITA result in accordance with an established cross-sectional incidence estimator^2^ and performance characteristics specified as a mean duration of recent infection = 130 days (95% CI: 118, 142), time cut-off = 1.0 year and residual proportion false recent = 0.0% (note that an assumption of FRR equal to 0.0% is explicitly not recommended in the 2011 WHO guidance;^16^ in addition, no uncertainty in the FRR estimate was accounted for in these analyses). No adjustment for subtype-related variation in MDRI was made. Survey weights were utilized for all estimates to account for the complex sampling design. The sample size of PHIA surveys is designed to provide subnational-level (e.g., provinces, regions) estimates of viral load suppression among people living with HIV (PLWH) aged 15 to 49 years with a 95% CI +/- 10% or less, which typically yield reasonably precise estimates of national-level HIV incidence among people aged 15 to 49 years. As a result, these surveys were able to generate HIV incidence estimates disaggregated by sex and high-level region, but not estimates that could be used to target HIV prevention or care to specific districts and/or key populations.

### Non-Incidence Surveillance Use Cases of HIV Recency Assays

Though there were 11 peer-reviewed studies reporting on non-incidence use cases identified across a 9-year review period, evidence on non-incidence use cases derived from routine HIV programme settings was scant. Four of these studies were from a single upper-income country (Ireland), three of which reported annual updates on approximately the same method of national implementation.

Beginning in 2016,^91,102^ Ireland’s Health Protection Surveillance Centre integrated recency testing combined with epidemiological data into national HIV surveillance to better monitor and inform HIV prevention interventions. In this system, normalised optical density (ODn) of new HIV diagnoses (measured using the Sedia^™^ HIV-1 Limiting Antigen-Avidity EIA) was linked with data captured in the national infectious disease reporting system. People with new diagnoses were classified as recent based on an ODn ≤ 1.5, unless epidemiological or clinical criteria (CD4 count <200 cells/mm^3^; viral load <400 copies/ml; the presence of AIDS-defining illness; prior ART use) indicated a probable false-recent result. During the pilot implementation of recency surveillance in Ireland in 2016, 448 of 508 (88.1%) new diagnoses nationwide were linked to a recency test result, with 12.5% of new diagnoses classified as recent. People who inject drugs had the highest proportion of recent infections (26.3% of new diagnoses were recent) and recent infection was significantly more likely with a concurrent sexually transmitted infection (aOR 2.59; 95% CI 1.04–6.45). However, data were incomplete for at least one RITA criterion in 48% of cases. Since then, further efforts have been made to improve completeness of the required epidemiological data, with national reports published for 2017^94^ and 2018.^95^ This example demonstrates the feasibility of integrating a RITA into routine surveillance, along with the challenges of identifying demographic sub-groups with ongoing HIV transmission when surveillance data are incomplete. However, it does not attempt to distinguish populations with high proportions of recent infections due to ongoing transmission from those experiencing improvements in testing frequency (which would lead to more diagnoses early in infection), and no details were available about potential systems modifications that resulted from this change to the surveillance strategy.

We identified two non-incidence, case surveillance reports from countries that used genetic testing combined with RITAs to better understand transmission dynamics.^90,92^ In one example, researchers in Belgium analysed HIV-1 *pol* sequences obtained through baseline drug resistance testing of patients newly diagnosed with HIV between 2013 and 2017.^92^ Information on genetic similarity was combined with demographic data and information on the recency of infection for 927 patients. They found that 48.3% of the patients were genetically linked to others, with 11.4% belonging to a pair and 36.9% involved in a cluster of ≥3 members. Patients of Belgian origin were more frequently involved in transmission clusters (49.7% compared to 15.3%) and diagnosed earlier (37.4% compared to 12.2%) than patients of Sub-Saharan African origin. Of the infections reported to be locally acquired, 69.5% were linked (14.1% paired and 55.4% in a cluster). Interestingly, equal proportions of early and late diagnosed individuals (59.9% and 52.4%, respectively) were involved in clusters, calling into question the added benefit of recency testing for molecular surveillance activities. The researchers argued that identification of a genetically linked individual for the majority of locally infected patients suggested a high rate of diagnosis in this population. However, frequent delays in diagnoses after infection increased opportunities for onward transmission, thus indicating that earlier diagnosis should be prioritized to protected HIV-uninfected members of sexual networks.

Finally, although this review only identified two pieces of evidence related to the *targeted prevention* use case, more evidence of using HIV recency assays for this purpose will likely emerge from the U.S. President’s Emergency Plan for AIDS Relief (PEPFAR) “TRACE” initiative (Tracking with Recency Assays to Control the Epidemic) in the near future. Beginning in Fiscal Year 2019, PEPFAR funded 16 countries (El Salvador, Eswatini, Ethiopia, Guatemala, Kenya, Lesotho, Malawi, Namibia, Nicaragua, Panama, Rwanda, Tanzania, Uganda, Vietnam, Zambia, Zimbabwe) who are nearing the 90–90–90 targets to introduce the TRACE initiative.^103^ Through TRACE, a lateral flow rapid recency assay is conducted as a supplementary test in routine HIV testing services and/or within HIV case surveillance to detect recent infection among newly diagnosed PLWH in all (or most) facility-and community-based testing sites in a country to drive prevention and care planning.

A variety of terms have been used to describe the analytic goals of TRACE programs including to identify, investigate, and intervene on; “areas with ongoing active transmission,” “hot-spots,” “sub-populations with high levels of HIV recency,” “clusters,” “pockets,” and “outbreaks.” In a 2018 viewpoint article promoting the TRACE initiative, Kim *et al*. argued that point-of-care recency assays used in conjunction with geographic data will allow jurisdictions to immediately hone in on *“hotspot”* locations and *“sub-populations with high levels of HIV recency,”* or to facilitate the identification and investigation of *“clusters”* of recent infections, triggering a public health response.^104^ PEPFAR’s 2019 annual report included narrative text stating that, “When used as an ancillary test in all those who are newly diagnosed with HIV, recency testing enables the identification of recent transmission *“pockets*.*”*^105^ More recently, the PEPFAR annual report for 2021 described using recency testing to identify where active transmission is occurring as HIV *“outbreak”* investigations.^106^ More precise and/or unified terms and indicators across all settings would improve interpretation and comparability across countries and regions. Furthermore, misidentification of clusters, hotspots and other imprecisely defined indicators through recency testing may result in misdirected or poorly designed prevention plans and missed opportunities for targeting limited resources.

## Limitations

There are several limitations to our systematic review. First, while the majority of the literature included in this review (74 of 90 pieces of evidence) came from the systematic review, additional pieces of evidence were submitted in response to the WHO country survey that were not identified during our systematic search; therefore, this portion of the systematic review is not reproducible and may suffer from selection bias. Second, given our search strategy many of the articles included in this review involved findings relevant to the performance of specific commercially available recency assays. However, some of those assays (e.g., the Sedia^™^ BED HIV-1 Incidence EIA) are technically available but no longer in wide use, due to inferior performance for HIV incidence estimation compared to other available assays. Further, some assays included in this review are not available in all countries globally. Third, as with all systematic reviews, our review process was imperfect and time-limited. Some meaningful literature was noted by our team after the review was complete,^107-110^ and has not been included to preserve fidelity to our pre-specified protocol.

## CONCLUSIONS

Despite widespread use of HIV recency assays for both HIV incidence estimation and non-incidence surveillance use cases, evidence on validated and accurate uses of recency assays for non-incidence surveillance remains weak. Further, more consistent and precise use of terminology is warranted, as is more rigorous validation of non-incidence indicators and methodologies to inform programme management. The evidence identified through this review will be used in a forthcoming technical guidance on the use of HIV recency assays for surveillance use cases, to be released by WHO and UNAIDS. This document is expected to help raise global awareness of benefits and pitfalls of the use of these assays for surveillance purposes, and set clear standards for their appropriate use. Based on the evidence identified through this review, this guidance will be able to provide strong updated recommendations about methods for population-level HIV incidence estimation. However, lack of evidence validating the accuracy and effectiveness of using recency assays for surveillance use cases other than incidence estimation will likely lead to less strong recommendations in those areas, and/or inability to make specific recommendations for use of recency assays for these purposes.

## Data Availability

Details of the search terms and results of this systematic review are available as supplemental material.

## AUTHORSHIP

SNF and LA conducted the systematic review and country survey, with advice and support from ADM and EG. SNF and ADM wrote the initial draft. KM, DL, SD, and MM provided funding and oversight of the project. All authors provided substantive revisions to the manuscript.

## ACKNOWLEDGMENTS

We acknowledge Virginia Fonner, Theresa Yeh, Cheryl Case Johnson, Anita Sands, and Rachel Clare Baggaley, who helped with the development and distribution of the global Call for Information from WHO member countries and participants of the WHO HIV Recency Testing Working Group. Thanks also to those who responded to this Call for Information, including WHO country office staff, researchers, public health surveillance staff, and HIV testing programme staff throughout the world.

## REFERENCES

1. Welte A, McWalter TA, Laeyendecker O, Hallett TB. Using tests for recent infection to estimate incidence: problems and prospects for HIV. Euro surveillance. 2010;15(24).

2. Kassanjee R, McWalter TA, Barnighausen T, Welte A. A new general biomarker-based incidence estimator. Epidemiology. 2012;23(5):721–8.

3. FIND. Target Product Profile for Tests for Recent HIV Infection. 2017 February.

4. Guy R, Gold J, Calleja JM, Kim AA, Parekh B, Busch M, et al. Accuracy of serological assays for detection of recent infection with HIV and estimation of population incidence: a systematic review. The Lancet infectious diseases. 2009;9(12):747–59.

5. Murphy G, Pilcher CD, Keating SM, Kassanjee R, Facente SN, Welte A, et al. Moving towards a reliable HIV incidence test - current status, resources available, future directions and challenges ahead. Epidemiology and infection. 2016:1–17.

6. Chaillon A, Le Vu S, Brunet S, Gras G, Bastides F, Bernard L, et al. Decreased specificity of an assay for recent infection in HIV-1-infected patients on highly active antiretroviral treatment: implications for incidence estimates. Clin Vaccine Immunol. 2012;19(8):1248–53.

7. Schlusser KE, Pilcher C, Kallas EG, Santos BR, Deeks SG, Facente S, et al. Comparison of cross-sectional HIV incidence assay results from dried blood spots and plasma. PloS one. 2017;12(2):e0172283.

8. Longosz AF, Serwadda D, Nalugoda F, Kigozi G, Franco V, Gray RH, et al. Impact of HIV subtype on performance of the limiting antigen-avidity enzyme immunoassay, the bio-rad avidity assay, and the BED capture immunoassay in Rakai, Uganda. AIDS research and human retroviruses. 2014;30(4):339–44.

9. Keating SM, Hanson D, Lebedeva M, Laeyendecker O, Ali-Napo NL, Owen SM, et al. Lower-sensitivity and avidity modifications of the vitros anti-HIV 1+2 assay for detection of recent HIV infections and incidence estimation. Journal of clinical microbiology. 2012;50(12):3968–76.

10. Laeyendecker O, Brookmeyer R, Oliver AE, Mullis CE, Eaton KP, Mueller AC, et al. Factors associated with incorrect identification of recent HIV infection using the BED capture immunoassay. AIDS research and human retroviruses. 2012;28(8):816–22.

11. Fogel JM, Piwowar-Manning E, Debevec B, Walsky T, Schlusser K, Laeyendecker O, et al. Brief Report: Impact of Early Antiretroviral Therapy on the Performance of HIV Rapid Tests and HIV Incidence Assays. Journal of acquired immune deficiency syndromes. 2017;75(4):426–30.

12. Klock E, Mwinnya G, Eller LA, Fernandez RE, Kibuuka H, Nitayaphan S, et al. Impact of Early Antiretroviral Treatment Initiation on Performance of Cross-Sectional Incidence Assays. AIDS research and human retroviruses. 2020;36(7):583–9.

13. World Health Organization. Meeting Report: WHO Working Group on HIV Incidence Measurement and Data Use. Boston: World Health Organization; 2018.

14. Gonese E, Kilmarx PH, van Schalkwyk C, Grebe E, Mutasa K, Ntozini R, et al. Evaluation of the Performance of Three Biomarker Assays for Recent HIV Infection Using a Well-Characterized HIV-1 Subtype C Incidence Cohort. AIDS research and human retroviruses. 2019;35(7):615–27.

15. Grebe E, Murphy G, Keating SM, Hampton D, Busch MP, Facente SN, et al. Impact of HIV-1 Subtype and Sex on Sedia Limiting Antigen Avidity Assay Performance. Conference on Retroviruses and Opportunistic Infections (CROI); Seattle, WA2019.

16. UNAIDS/WHO Working Group on Global HIV/AIDS and STI Surveillance. When and How to Use Assays for Recent Infection to Estimate HIV Incidence at a Population Level. Geneva; 2011.

17. World Health Organization (WHO), Joint United Nations Programme on HIV/AIDS (UNAIDS). WHO/UNAIDS Technical Update on HIV Incidence Assays for Surveillance and Epidemic Monitoring. Geneva; 2013 May 30.

18. Joint United Nations Programme on HIV/AIDS (UNAIDS), World Health Organization (WHO). Technical Update on HIV Incidence Assays for Surveillance and Monitoring Purposes. Geneva; 2015.

19. Global HIV Strategic Information Working Group. Recent Infection Testing Algorithm Technical Update: Applications for HIV surveillance and programme monitoring. Geneva; 2018.

20. World Health O, Unaids. WHO Working Group on HIV incidence assays: estimating HIV incidence using HIV case surveillance: meeting report, Glion, Switzerland, 10–11 December 2015. Geneva: World Health Organization; 2017 2017.

21. Mammone A, Pezzotti P, Angeletti C, Orchi N, Carboni A, Navarra A, et al. HIV incidence estimate combining HIV/AIDS surveillance, testing history information and HIV test to identify recent infections in Lazio, Italy. BMC Infect Dis. 2012;12:65.

22. Zhu Q, Wang Y, Liu J, Duan X, Chen M, Yang J, et al. Identifying major drivers of incident HIV infection using recent infection testing algorithms (RITAs) to precisely inform targeted prevention. International journal of infectious diseases:IJID:official publication of the International Society for Infectious Diseases. 2020;101:131–7.

23. Laeyendecker O, Brookmeyer R, Mullis CE, Donnell D, Lingappa J, Celum C, et al. Specificity of four laboratory approaches for cross-sectional HIV incidence determination: analysis of samples from adults with known nonrecent HIV infection from five African countries. AIDS research and human retroviruses. 2012;28(10):1177–83.

24. Cousins MM, Konikoff J, Sabin D, Khaki L, Longosz AF, Laeyendecker O, et al. A comparison of two measures of HIV diversity in multi-assay algorithms for HIV incidence estimation. PloS one. 2014;9(6):e101043.

25. Karatzas-Delgado EF, Ruiz-González V, García-Cisneros S, Olamendi-Portugal ML, Herrera-Ortiz A, López-Gatell H, et al. Evaluation of an HIV recent infection testing algorithm with serological assays among men who have sex with men in Mexico. Journal of infection and public health. 2020;13(4):509–13.

26. Konikoff J, Brookmeyer R, Longosz AF, Cousins MM, Celum C, Buchbinder SP, et al. Performance of a limiting-antigen avidity enzyme immunoassay for cross-sectional estimation of HIV incidence in the United States. PloS one. 2013;8(12):e82772.

27. Kim AA, Rehle T. Short Communication: Assessing Estimates of HIV Incidence with a Recent Infection Testing Algorithm That Includes Viral Load Testing and Exposure to Antiretroviral Therapy. AIDS research and human retroviruses. 2018;34(10):863–6.

28. Shah NS, Duong YTL.LV, Tuan NA, Parekh BS, Ha HTT, et al. Estimating False-Recent Classification for the Limiting-Antigen Avidity EIA and BED-Capture Enzyme Immunoassay in Vietnam: Implications for HIV-1 Incidence Estimates. AIDS research and human retroviruses. 2017;33(6):546–54.

29. Keating SM, Kassanjee R, Lebedeva M, Facente SN, MacArthur JC, Grebe E, et al. Performance of the Bio-Rad Geenius HIV1/2 Supplemental Assay in Detecting “Recent” HIV Infection and Calculating Population Incidence. Journal of acquired immune deficiency syndromes. 2016;73(5):581–8.

30. Duong YT, Kassanjee R, Welte A, Morgan M, De A, Dobbs T, et al. Recalibration of the limiting antigen avidity EIA to determine mean duration of recent infection in divergent HIV-1 subtypes. PloS one. 2015;10(2):e0114947.

31. McNicholl JM, McDougal JS, Wasinrapee P, Branson BM, Martin M, Tappero JW, et al. Assessment of BED HIV-1 incidence assay in seroconverter cohorts: effect of individuals with long-term infection and importance of stable incidence. PloS one. 2011;6(3):e14748.

32. Hargrove J, van Schalkwyk C, Eastwood H. BED estimates of HIV incidence: resolving the differences, making things simpler. PloS one. 2012;7(1):e29736.

33. Xu Y, Laeyendecker O, Wang R. Cross-sectional human immunodeficiency virus incidence estimation accounting for heterogeneity across communities. Biometrics. 2019;75(3):1017–28.

34. Duong YT, Qiu M, De AK, Jackson K, Dobbs T, Kim AA, et al. Detection of recent HIV-1 infection using a new limiting-antigen avidity assay: potential for HIV-1 incidence estimates and avidity maturation studies. PloS one. 2012;7(3):e33328.

35. Parekh BS, Hanson DL, Hargrove J, Branson B, Green T, Dobbs T, et al. Determination of mean recency period for estimation of HIV type 1 Incidence with the BED-capture EIA in persons infected with diverse subtypes. AIDS research and human retroviruses. 2011;27(3):265–73.

36. Kirkpatrick AR, Patel EU, Celum CL, Moore RD, Blankson JN, Mehta SH, et al. Development and Evaluation of a Modified Fourth-Generation Human Immunodeficiency Virus Enzyme Immunoassay for Cross-Sectional Incidence Estimation in Clade B Populations. AIDS research and human retroviruses. 2016;32(8):756–62.

37. Keating SM, Rountree W, Grebe E, Pappas AL, Stone M, Hampton D, et al. Development of an international external quality assurance program for HIV-1 incidence using the Limiting Antigen Avidity assay. PloS one. 2019;14(9):e0222290.

38. Braunstein SL, Nash D, Kim AA, Ford K, Mwambarangwe L, Ingabire CM, et al. Dual testing algorithm of BED-CEIA and AxSYM Avidity Index assays performs best in identifying recent HIV infection in a sample of Rwandan sex workers. PloS one. 2011;6(4):e18402.

39. Kim AA, Hallett T, Stover J, Gouws E, Musinguzi J, Mureithi PK, et al. Estimating HIV incidence among adults in Kenya and Uganda: a systematic comparison of multiple methods. PloS one. 2011;6(3):e17535.

40. Mastro TD, Kim AA, Hallett T, Rehle T, Welte A, Laeyendecker O, et al. Estimating HIV Incidence in Populations Using Tests for Recent Infection: Issues, Challenges and the Way Forward. Journal of HIV AIDS surveillance & epidemiology. 2010;2(1):1–14.

41. Brookmeyer R, Konikoff J, Laeyendecker O, Eshleman SH. Estimation of HIV incidence using multiple biomarkers. American journal of epidemiology. 2013;177(3):264–72.

42. Laeyendecker O, Brookmeyer R, Cousins MM, Mullis CE, Konikoff J, Donnell D, et al. HIV incidence determination in the United States: a multiassay approach. The Journal of infectious diseases. 2013;207(2):232–9.

43. Vermeulen M, Chowdhury D, Swanevelder R, Grebe E, Brambilla D, Jentsch U, et al. HIV incidence in South African blood donors from 2012 to 2016: a comparison of estimation methods. Vox Sanguinis. 2021;116(1):71–80.

44. Longosz AF, Morrison CS, Chen PL, Arts E, Nankya I, Salata RA, et al. Immune responses in Ugandan women infected with subtypes A and D HIV using the BED capture immunoassay and an antibody avidity assay. Journal of acquired immune deficiency syndromes. 2014;65(4):390–6.

45. Hauser A, Santos-Hoevener C, Meixenberger K, Zimmermann R, Somogyi S, Fiedler S, et al. Improved testing of recent HIV-1 infections with the BioRad avidity assay compared to the limiting antigen avidity assay and BED Capture enzyme immunoassay: evaluation using reference sample panels from the German Seroconverter Cohort. PloS one. 2014;9(6):e98038.

46. Bao L, Ye J, Hallett TB. Incorporating incidence information within the UNAIDS Estimation and Projection Package framework: a study based on simulated incidence assay data. Aids. 2014;28 Suppl 4(4):S515–22.

47. Grebe E, Welte A, Hall J, Keating SM, Facente SN, Marson K, et al. Infection Staging and Incidence Surveillance Applications of High Dynamic Range Diagnostic Immuno-Assay Platforms. Journal of acquired immune deficiency syndromes. 2017;76(5):547–55.

48. Mahiane SG, Fiamma A, Auvert B. Mixture models for calibrating the BED for HIV incidence testing. Statistics in medicine. 2014;33(10):1767–83.

49. Sempa JB, Welte A, Busch MP, Hall J, Hampton D, Facente SN, et al. Performance comparison of the Maxim and Sedia Limiting Antigen Avidity assays for HIV incidence surveillance. PloS one. 2019;14(7):e0220345.

50. Serhir B, Hamel D, Doualla-Bell F, Routy JP, Beaulac SN, Legault M, et al. Performance of Bio-Rad and Limiting Antigen Avidity Assays in Detecting Recent HIV Infections Using the Quebec Primary HIV-1 Infection Cohort. PloS one. 2016;11(5):e0156023.

51. Schlusser KE, Konikoff J, Kirkpatrick AR, Morrison C, Chipato T, Chen PL, et al. Short Communication: Comparison of Maxim and Sedia Limiting Antigen Assay Performance for Measuring HIV Incidence. AIDS research and human retroviruses. 2017;33(6):555–7.

52. Yu L, Laeyendecker O, Wendel SK, Liang F, Liu W, Wang X, et al. Short Communication: Low False Recent Rate of Limiting-Antigen Avidity Assay Among Long-Term Infected Subjects from Guangxi, China. AIDS research and human retroviruses. 2015;31(12):1247–9.

53. Schüpbach J, Gebhardt MD, Tomasik Z, Niederhauser C, Yerly S, Bürgisser P, et al. Assessment of recent HIV-1 infection by a line immunoassay for HIV-1/2 confirmation. PLoS Med. 2007;4(12):e343.

54. Huik K, Soodla P, Pauskar M, Owen SM, Luo W, Murphy G, et al. The concordance of the limiting antigen and the Bio-Rad avidity assays in persons from Estonia infected mainly with HIV-1 CRF06_cpx. PloS one. 2019;14(5):e0217048.

55. Kassanjee R, Pilcher CD, Busch MP, Murphy G, Facente SN, Keating SM, et al. Viral load criteria and threshold optimization to improve HIV incidence assay characteristics. Aids. 2016;30(15):2361–71.

56. Kassanjee R, Pilcher CD, Keating SM, Facente SN, McKinney E, Price MA, et al. Independent assessment of candidate HIV incidence assays on specimens in the CEPHIA repository. Aids. 2014;28(16):2439–49.

57. Huerga H, Shiferie F, Grebe E, Giuliani R, Farhat JB, Van-Cutsem G, et al. A comparison of self-report and antiretroviral detection to inform estimates of antiretroviral therapy coverage, viral load suppression and HIV incidence in Kwazulu-Natal, South Africa. BMC Infect Dis. 2017;17(1):653.

58. Rehle T, Johnson L, Hallett T, Mahy M, Kim A, Odido H, et al. A Comparison of South African National HIV Incidence Estimates: A Critical Appraisal of Different Methods. PloS one. 2015;10(7):e0133255.

59. Grebe E, Welte A, Hall J, Busch MP, Facente SN, Keating S, et al. Recency staging of HIV infections through routine diagnostic testing [Poster]. Conference on Retroviruses and Opportunistic Infections (CROI); Boston, MA, 2017.

60. Ramos EM, Ortega J, Daza G, Namking Y, Harb S, Dragavon J, et al. Use of the Sample-to-Cutoff Ratio (S/CO) to Identify Recency of HIV-1 Infection [Poster]. Conference on Retroviruses and Opportunistic Infections (CROI); Seattle, WA, 2015.

61. Grebe E, Vermeulen M, Brits T, Swanevelder R, Jacobs G, Busch MP, et al. Performance Validation of the Sedia HIV-1 Limiting Antigen (LAg)-Avidity EIA in South African Blood Donors [Poster]. Conference on Retroviruses and Opportunistic Infections (CROI); Boston, MA, 2018.

62. Grebe E, Facente SN, Owen R, Hampton D, Cheng C, Sharma U, et al. Independent Assessment of the Sedia Asante HIV-1 Rapid Recency Assay. HIV Diagnostics Conference; Atlanta, GA, 2019.

63. Laeyendecker O, Gray RH, Grabowski MK, Reynolds SJ, Ndyanabo A, Ssekasanvu J, et al. Validation of the Limiting Antigen Avidity Assay to Estimate Level and Trends in HIV Incidence in an A/D Epidemic in Rakai, Uganda. AIDS research and human retroviruses. 2019;35(4):364–7.

64. Guy RJ, Breschkin AM, Keenan CM, Catton MG, Enriquez AM, Hellard ME. Improving HIV surveillance in Victoria: the role of the “detuned” enzyme immunoassay. Journal of acquired immune deficiency syndromes. 2005;38(4):495–9.

65. El-Hayek C, Breschkin A, Nicholson S, Bergeri I, Hellard ME. Does Using a Bed Enzyme Immunoassay Test Enhance Current HIV Surveillance Practices? [Poster]. XVIII International AIDS Conference; Vienna, Austria, 2010.

66. Verhofstede C, Fransen K, Van Den Heuvel A, Van Laethem K, Ruelle J, Vancutsem E, et al. Decision tree for accurate infection timing in individuals newly diagnosed with HIV-1 infection. BMC Infect Dis. 2017;17(1):738.

67. Hassan J, Moran J, Murphy G, Mason O, Connell J, De Gascun C. Discrimination between recent and non-recent HIV infections using routine diagnostic serological assays. Medical microbiology and immunology. 2019.

68. Grebe E, Facente SN, Hampton D, Cheng C, Owen R, Keating SM, et al. Evaluation of the Asante HIV-1 Rapid Recency Assay. San Francisco, CA; 2019.

69. Duong YT, Dobbs T, Mavengere Y, Manjengwa J, Rottinghaus E, Saito S, et al. Field Validation of Limiting-Antigen Avidity Enzyme Immunoassay to Estimate HIV-1 Incidence in Cross-Sectional Survey in Swaziland. AIDS research and human retroviruses. 2019;35(10):896–905.

70. Yufenyuy E, Detorio M, Tan X, Shanmugam V, Dobbs T, Kim A, et al. Evaluation of Rapid Tests for Recent HIV Infection: Implications for Real-time Surveillance and Epidemic Control [Poster]. The 10th IAS Conference on HIV Science; Mexico City, Mexico, 2019.

71. Rakai Health Sciences Program, Uganda Virus Research Institute (UVRI) - HIV Reference Laboratory, MRC/UVRI, London School of Hygiene & Tropical Medicine Uganda Research Unit. Validation of the Asante HIV-1 Rapid Recency Assay for recent HIV-1 infection detection in Uganda. Rakai: Ministry of Health, Uganda; 2020.

72. Aghaizu A, Tosswill J, De Angelis D, Ward H, Hughes G, Murphy G, et al. HIV incidence among sexual health clinic attendees in England: First estimates for black African heterosexuals using a biomarker, 2009-2013. PloS one. 2018;13(6):e0197939.

73. Garrett N, Lattimore S, Gilbart V, Aghaizu A, Mensah G, Tosswill J, et al. The Recent Infection Testing Algorithm (RITA) in clinical practice: a survey of HIV clinicians in England and Northern Ireland. HIV medicine. 2012;13(7):444–7.

74. Kim AA, McDougal JS, Hargrove J, Rehle T, Pillay-Van Wyk V, Puren A, et al. Evaluating the BED capture enzyme immunoassay to estimate HIV incidence among adults in three countries in sub-Saharan Africa. AIDS research and human retroviruses. 2010;26(10):1051–61.

75. Ministry of Health U. Uganda Population-based HIV Impact Assessment (UPHIA) 2016-2017: Final Report. Kampala: Ministry of Health; 2019 July.

76. Ministry of Health M. Malawi Population-Based HIV Impact Assessment (MPHIA), 2015-2016: Final Report. Lilongwe: Ministry of Health; 2018 October.

77. Ministry of Health and Child Care (MOHCC) Z. Zimbabwe Population-based HIV Impact Assessment (ZIMPHIA) 2015-2016: Final Report. Harare: MOHCC; 2019 August.

78. Ministry of Health L, Centers for Disease Control and Prevention (CDC), ICAP at Columbia University. Lesotho Population-based HIV Impact Assessment (LePHIA) 2016-2017: Final Report. Maseru, Lesotho, Atlanta, Georgia, and New York, New York: Ministry of Health, CDC, and ICAP; 2019 September.

79. Government of the Kingdom of Eswatini. Swaziland HIV Incidence Measurement Survey 2 (SHIMS2) 2016-2017: Final Report. Mbabane: Government of the Kingdom of Eswantini; 2019 April.

80. Tanzania Commission for AIDS (TACAIDS), Zanzibar AIDS Commission (ZAC). Tanzania HIV Impact Survey (THIS) 2016-2017: Final Report. Dar es Salaam: TACAIDS, ZAC; 2018 December.

81. Ministry of Health Z. Zambia Population-based HIV Impact Assessment (ZAMPHIA) 2016: Final Report. Lusaka: Ministry of Health; 2019 February.

82. Ministry of Health and Social Services (MoHSS) N. Namibia Population-based HIV Impact Assessment (NAMPHIA) 2017: Final Report. Windhoek: MoHSS, Namibia; 2019 November.

83. Ethiopian Public Health Institute (EPHI). Ethiopia Population-based HIV Impact Assessment (EPHIA) 2017-2018: Final Report. Addis Ababa: EPHI; 2020 August.

84. Rwanda Biomedical Center (RBC). Rwanda Population-based HIV Impact Assessment (RPHIA) 2018-2019: Final Report. Kigali: RBC; 2020 September.

85. Simbayi LC, Zuma K, Zungu N, Moyo S, Marinda E, Jooste S, et al. South African National HIV Prevalence, Incidence, Behaviour and Communication Survey, 2017. Cape Town:: HSRC Press; 2019.

86. Soodla P, Simmons R, Huik K, Pauskar M, Jõgeda EL, Rajasaar H, et al. HIV incidence in the Estonian population in 2013 determined using the HIV-1 limiting antigen avidity assay. HIV medicine. 2018;19(1):33–41.

87. Grebe E, Busch MP, Notari EP, Bruhn R, Quiner C, Hindes D, et al. HIV incidence in US first-time blood donors and transfusion risk with a 12-month deferral for men who have sex with men. Blood. 2020;136(11):1359–67.

88. de Oliveira Garcia Mateos S, Preiss L, Gonçalez TT, Di Lorenzo Oliveira C, Grebe E, Di Germanio C, et al. 10-year analysis of human immunodeficiency virus incidence in first-time and repeat donors in Brazil. Vox Sang. 2021;116(2):207–16.

89. Sexton CJ, Costenbader EC, Vinh DT, Chen PL, Hoang TV, Lan NT, et al. Correlation of prospective and cross-sectional measures of HIV type 1 incidence in a higher-risk cohort in Ho Chi Minh City, Vietnam. AIDS research and human retroviruses. 2012;28(8):866–73.

90. Otecko N, Inzaule S, Odhiambo C, Otieno G, Opollo V, Morwabe A, et al. Viral and Host Characteristics of Recent and Established HIV-1 Infections in Kisumu based on a Multiassay Approach. Sci Rep. 2016;6:37964.

91. Robinson E, Moran J, O’Donnell K, Hassan J, Tuite H, Ennis O, et al. Integration of a recent infection testing algorithm into HIV surveillance in Ireland: improving HIV knowledge to target prevention. Epidemiology and infection. 2019;147:e136.

92. Verhofstede C, Mortier V, Dauwe K, Callens S, Deblonde J, Dessilly G, et al. Exploring HIV-1 Transmission Dynamics by Combining Phylogenetic Analysis and Infection Timing. Viruses. 2019;11(12).

93. Giese C, Igoe D, Gibbons Z, Hurley C, Stokes S, McNamara S, et al. Injection of new psychoactive substance snow blow associated with recently acquired HIV infections among homeless people who inject drugs in Dublin, Ireland, 2015. Euro surveillance. 2015;20(40).

94. Health Service Executive (HSE) Health Protection Surveillance Centre (HPSC). Monitoring Recent HIV Infection in Ireland, 2017. Dublin: HSE HPSC; 2019.

95. HSE HPSC. Monitoring Recent HIV Infection in Ireland, 2018. Dublin: HSE HPSC; 2020.

96. The Monitoring Recently Acquired HIV Infection Group. Integration of recent infection monitoring into national HIV surveillance: 2016 results. Dublin, Ireland; 2018.

97. Aghaizu A, Murphy G, Tosswill J, DeAngelis D, Charlett A, Gill ON, et al. Recent infection testing algorithm (RITA) applied to new HIV diagnoses in England, Wales and Northern Ireland, 2009 to 2011. Euro surveillance. 2014;19(2).

98. Rice BD, de Wit M, Welty S, Risher K, Cowan FM, Murphy G, et al. Can HIV recent infection surveillance help us better understand where primary prevention efforts should be targeted? Results of three pilots integrating a recent infection testing algorithm into routine programme activities in Kenya and Zimbabwe. Journal of the International AIDS Society. 2020;23 Suppl 3(Suppl 3):e25513.

99. UNAIDS/WHO Working Group on Global HIV/AIDS and STI Surveillance. Monitoring HIV Impact Using Population-Based Surveys. Geneva; 2015.

100. ICAP, Columbia University. Population-based HIV Impact Assessment: Guiding the Global HIV Response 2021 [Available from: https://phia.icap.columbia.edu/about.

101. ICAP. PHIA Project Countries. New York, NY: Columbia University; 2019 [Available from: https://phia.icap.columbia.edu/countries-overview/.

102. HSE HPSC. Monitoring Recent HIV Infection in Ireland, 2016. Dublin: HSE HPSC; 2018.

103. The TRACE Inititiave. TRACE - Tracking with Recency Assays to Control the Epidemic 2021 [Available from: https://trace-recency.org.

104. Kim AA, Behel S, Northbrook S, Parekh BS. Tracking with recency assays to control the epidemic: real-time HIV surveillance and public health response. Aids. 2019;33(9):1527–9.

105. President’s Emergency Plan for AIDS Relief (PEPFAR). PEPFAR 2019 Annual Report to Congress. Washington, D.C.: PEPFAR; 2019.

106. PEPFAR. PEPFAR 2021 Annual Report to Congress. Washington, D.C.: PEPFAR; 2021.

107. Galiwango RM, Ssuuna C, Kaleebu P, Kigozi G, Kagaayi J, Nakigozi G, et al. Validation of the Asante HIV-1 Rapid Recency Assay for Detection of Recent HIV-1 Infections in Uganda. AIDS research and human retroviruses. 2021.

108. Hargrove JW, Humphrey JH, Mutasa K, Parekh BS, McDougal JS, Ntozini R, et al. Improved HIV-1 incidence estimates using the BED capture enzyme immunoassay. Aids. 2008;22(4):511–8.

109. Ministry of Health M. Estimating HIV Incidence and Detecting Recent Infection Among Pregnant Adolescent Girls and Young Women in Malawi, 2017-2018: Final Report. Lilongwe: Ministry of Health; 2019 October

110. Welty S, Motoku J, Muriithi C, Rice B, de Wit M, Ashanda B, et al. Brief Report: Recent HIV Infection Surveillance in Routine HIV Testing in Nairobi, Kenya: A Feasibility Study. Journal of Acquired Immune Deficiency Syndromes. 2020;84(1):5–9.

111. Cohen JF, Korevaar DA, Altman DG, Bruns DE, Gatsonis CA, Hooft L, et al. STARD 2015 guidelines for reporting diagnostic accuracy studies: explanation and elaboration. BMJ Open. 2016;6(11):e012799.

