## Supplemental Tables S1 - S5 for "Use of HIV Recency Assays for HIV Incidence Estimation and Non-Incidence Surveillance Use Cases: A systematic review"

**Table S1.** Search sets and terms used for title, abstract, & MeSH terms/author keyword searches.

| **Strategy 1** | | |
| --- | --- | --- |
| **Date** | **Criterion 1** | **Criterion 2** |
| Jan 1, 2010 to date of search | HIV  AND  recency assay or  incidence assay or  test for recent infection or  TRI or  RTRI | performance or  false recent rate or  false recent or  proportion false recent or  FRR or  mean duration of recent infection or  MDRI |
| **Strategy 2** | | |
| **Date** | **Criterion 1** | **Criterion 2** |
| Jan 1, 2010 to date of search | HIV  AND  recent infection testing algorithm or  RITA or  recency or  recent infection or  recent HIV infection | incidence estimation or  incidence estimate or  incidence estimator or  hotspot or  cluster or  case surveillance or  case-based surveillance or  mapping |

**Table S2.** Search code.

| MEDLINE (PubMed) | **1^st^ Search: Focus on assay performance**  2010/01/01:3000/12/31[Date - Publication] AND (HIV[Title/Abstract] OR HIV[MeSH Terms]) AND ("recency assay"[Title/Abstract] OR "incidence assay"[Title/Abstract] OR "recency assay"[MeSH Terms] OR "incidence assay"[MeSH Terms] OR "test for recent infection"[Title/Abstract] OR TRI[Title/Abstract] OR RTRI[Title/Abstract]) AND (performance[Title/Abstract] OR performance[MeSH Terms] OR "false recent rate"[Title/Abstract] OR "false recent"[Title/Abstract] OR "proportion false recent"[Title/Abstract] OR (FRR[Title/Abstract] OR "mean duration of recent infection"[Title/Abstract] OR MDRI[Title/Abstract])  **2^nd^ Search:** **Focus on the use of recency testing, with special attention to variations in assays, settings, and methods of analysis for calculating HIV incidence estimates**  2010/01/01:3000/12/31[Date - Publication] AND ("HIV"[Title/Abstract] OR "HIV"[MeSH Terms]) AND ("recent infection testing algorithm"[Title/Abstract] OR "RITA"[Title/Abstract] OR "recency"[Title/Abstract] OR "recent infection"[Title/Abstract] OR "recent HIV infection"[Title/Abstract]) AND ("incidence estimat*"[Title/Abstract] OR hotspot[Title/Abstract] OR cluster[Title/Abstract] OR "case surveillance"[Title/Abstract] OR "case-based surveillance"[Title/Abstract] OR "mapping"[Title/Abstract]) |
| --- | --- |
| Web of Science | **1^st^ Search: Focus on assay performance**  (TI=(HIV AND ('recency assay' or 'incidence assay' or 'test for recent infection' or TRI or RTRI)) OR AB=(HIV AND ('recency assay' or 'incidence assay' or 'test for recent infection' or TRI or RTRI)) OR AK=(HIV AND ('recency assay' or 'incidence assay' or 'test for recent infection' or TRI or RTRI))) AND (TI=('performance' or 'false recent rate' or 'false recent' or 'proportion false recent' or FRR or 'mean duration of recent infection' or MDRI) OR AB=('performance' or 'false recent rate' or 'false recent' or 'proportion false recent' or FRR or 'mean duration of recent infection' or MDRI) OR AK=('performance' or 'false recent rate' or 'false recent' or 'proportion false recent' or FRR or 'mean duration of recent infection' or MDRI)) [IC Timespan = 2010 to 2021]  **2^nd^ Search:** **Focus on the use of recency testing, with special attention to variations in assays, settings, and methods of analysis for calculating HIV incidence estimates**  (TI=(HIV AND ('recent infection testing algorithm' or RITA or recency or 'recent infection' or 'recent HIV infection')) OR AB=(HIV AND ('recent infection testing algorithm' or RITA or recency or 'recent infection' or 'recent HIV infection')) OR AK=(HIV AND ('recent infection testing algorithm' or RITA or recency or 'recent infection' or 'recent HIV infection'))) AND (TI=('incidence estimate*' or hotspot or cluster or 'case surveillance' or 'case-based surveillance' or mapping) OR AB=('incidence estimate*' or hotspot or cluster or 'case surveillance' or 'case-based surveillance' or mapping) OR AK=('incidence estimate*' or hotspot or cluster or 'case surveillance' or 'case-based surveillance' or mapping)) [IC Timespan = 2010 to 2021] |

**Table S3.** Websites searched for eligible grey literature during the review.

| **Organization/search engine** | **Website** |
| --- | --- |
| Google scholar | <https://scholar.google.com/> |
| International AIDS Society | <https://www.iasociety.org/> |
| International AIDS Society conference on HIV Science | <https://www.ias2021.org/> |
| International AIDS Conferences from 2010-2020 | e.g., <https://www.aids2020.org/> |
| International AIDS Society conference on HIV Research for Prevention | <https://www.hivr4p.org/> |
| Conference on Retroviruses and Opportunistic Infections (CROI) | <https://www.croiconference.org/> |
| HIV Diagnostics Conference | <http://hivtestingconference.org/> |
| Measurement and Surveillance of HIV epidemics (MeSH Consortium) | <https://mesh-consortium.org.uk/> |

**Table S4**. Sources identified during a systematic review of the literature (as described in the Methods section; Section 1.5) are organized below. Sources are ordered by 1) literature type (peer-reviewed vs. grey), then 2) strength of evidence (highest to lowest), and then 3) last name of the first author (alphabetical).

| Source | Year | Topic | Setting | Assay(s) | Strength^[1]^ |
| --- | --- | --- | --- | --- | --- |
| Bao L, Ye J, Hallett TB. Incorporating incidence information within the UNAIDS Estimation and Projection Package framework: a study based on simulated incidence assay data. Aids. 2014;28 Suppl 4(4):S515-22. | 2014 | incidence estimation method | n/a | n/a | 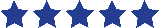 |
| Braunstein SL, Nash D, Kim AA, Ford K, Mwambarangwe L, Ingabire CM, et al. Dual testing algorithm of BED-CEIA and AxSYM Avidity Index assays performs best in identifying recent HIV infection in a sample of Rwandan sex workers. PLoS One. 2011;6(4):e18402. | 2011 | algorithm performance | Cross-sectional survey of Rwandan sex workers | BED (and AxSYM Avidity Index) | 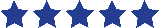 |
| Brookmeyer R, Konikoff J, Laeyendecker O, Eshleman SH. Estimation of HIV incidence using multiple biomarkers. Am J Epidemiol. 2013;177(3):264-72. | 2013 | incidence estimation method | US longitudinal cohorts | n/a | 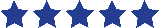 |
| Cousins MM, Konikoff J, Sabin D, Khaki L, Longosz AF, Laeyendecker O, et al. A comparison of two measures of HIV diversity in multi-assay algorithms for HIV incidence estimation. PLoS One. 2014;9(6):e101043. | 2014 | algorithm performance | US-based large clinical trials | Sedia LAg and BioRad Avidity | 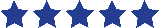 |
| Duong YT, Kassanjee R, Welte A, Morgan M, De A, Dobbs T, et al. Recalibration of the limiting antigen avidity EIA to determine mean duration of recent infection in divergent HIV-1 subtypes. PLoS One. 2015;10(2):e0114947. | 2015 | assay performance | Misc. global specimens of varying subtypes | Sedia LAg | 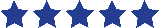 |
| Duong YT, Qiu M, De AK, Jackson K, Dobbs T, Kim AA, et al. Detection of recent HIV-1 infection using a new limiting-antigen avidity assay: potential for HIV-1 incidence estimates and avidity maturation studies. PLoS One. 2012;7(3):e33328. | 2012 | assay performance | n/a | BED and LAg (before commercial production) | 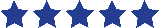 |
| Fogel JM, Piwowar-Manning E, Debevec B, Walsky T, Schlusser K, Laeyendecker O, et al. Brief Report: Impact of Early Antiretroviral Therapy on the Performance of HIV Rapid Tests and HIV Incidence Assays. J Acquir Immune Defic Syndr. 2017;75(4):426-30 | 2017 | assay performance | HPTN 052 in Malawi | Sedia LAg and BioRad Avidity | 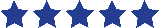 |
| Grebe E, Welte A, Hall J, Keating SM, Facente SN, Marson K, et al. Infection Staging and Incidence Surveillance Applications of High Dynamic Range Diagnostic Immuno-Assay Platforms. J Acquir Immune Defic Syndr. 2017;76(5):547-55. | 2017 | assay performance | CEHIA evaluation panel | Sedia LAg, Ortho VITROS, ARCHITECT | 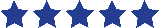 |
| Guy R, Gold J, Calleja JM, Kim AA, Parekh B, Busch M, et al. Accuracy of serological assays for detection of recent infection with HIV and estimation of population incidence: a systematic review. Lancet Infect Dis. 2009;9(12):747-59. | 2009 | assay performance | Systematic review | 13 assays | 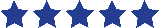 |
| Hargrove J, van Schalkwyk C, Eastwood H. BED estimates of HIV incidence: resolving the differences, making things simpler. PLoS One. 2012;7(1):e29736. | 2012 | incidence estimation method | n/a | BED | 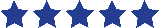 |
| Hargrove JW, Humphrey JH, Mutasa K, Parekh BS, McDougal JS, Ntozini R, et al. Improved HIV-1 incidence estimates using the BED capture enzyme immunoassay. Aids. 2008;22(4):511-8.^[2]^ | 2008 | assay performance | Postpartum women in Zimbabwe | BED | 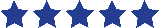 |
| Huerga H, Shiferie F, Grebe E, Giuliani R, Farhat JB, Van-Cutsem G, et al. A comparison of self-report and antiretroviral detection to inform estimates of antiretroviral therapy coverage, viral load suppression and HIV incidence in Kwazulu-Natal, South Africa. BMC Infect Dis. 2017;17(1):653. | 2017 | algorithm performance | Cross-sectional survey in Kwa-Zulu-Natal, South Africa | n/a | 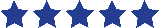 |
| Karatzas-Delgado EF, Ruiz-González V, García-Cisneros S, Olamendi-Portugal ML, Herrera-Ortiz A, López-Gatell H, et al. Evaluation of an HIV recent infection testing algorithm with serological assays among men who have sex with men in Mexico. J Infect Public Health. 2020;13(4):509-13. | 2019 | algorithm performance | A serological study of MSM in Mexico | BED and Maxim LAg | 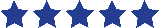 |
| Kassanjee R, Pilcher CD, Busch MP, Murphy G, Facente, SN, Keating SM, Mckinney E, Marson K, Price MA, Martin JN, Little SJ, Hecht FM, Kallas EG, Welte A, Consortium for the Evaluation and Performance of HIV Incidence Assays (CEPHIA). Viral load criteria and threshold optimization to improve HIV incidence assay characteristics*.* AIDS, 2016. 30(15): p. 2361-71. | 2016 | incidence estimation method | CEPHIA evaluation panels | 7 assays | 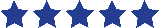 |
| Kassanjee R, Pilcher CD, Keating SM, Facente SN, McKinney E, Price MA, et al. Independent assessment of candidate HIV incidence assays on specimens in the CEPHIA repository. Aids. 2014;28(16):2439-49. | 2014 | assay performance | CEPHIA evaluation panels | Sedia LAg, BED, LS-Vitros, Vitros Avidity, BioRad Avidity | 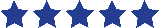 |
| Keating SM, Kassanjee R, Lebedeva M, Facente SN, MacArthur JC, Grebe E, Murphy G, Welte A, Martin JN, Little S, Price MA, Kallas EG, Busch MP, Pilcher CD. Performance of the Bio-Rad Geenius HIV1/2 Supplemental Assay in Detecting "Recent" HIV Infection and Calculating Population Incidence. J Acquir Immune Defic Syndr, 2016. 73(5): p. 581-588. | 2016 | assay performance | CEPHIA evaluation panels | BioRad Geenius | 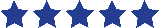 |
| Keating SM, Hanson D, Lebedeva M, Laeyendecker O, Ali-Napo NL, Owen SM, et al. Lower-sensitivity and avidity modifications of the vitros anti-HIV 1+2 assay for detection of recent HIV infections and incidence estimation. J Clin Microbiol. 2012;50(12):3968-76. | 2012 | assay performance | Seroconversion panels | Ortho VITROS (LS and Avidity) | 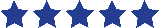 |
| Kim AA, Parekh BS, Umuro M, Galgalo T, Bunnell R, Makokha E, et al. Identifying Risk Factors for Recent HIV Infection in Kenya Using a Recent Infection Testing Algorithm: Results from a Nationally Representative Population-Based Survey. PLoS One. 2016;11(5):e0155498. | 2016 | field use of recency assays | 2007 Kenya AIDS Indicator Survey | Sedia LAg | 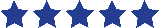 |
| Kim AA, Hallett T, Stover J, Gouws E, Musinguzi J, Mureithi PK, et al. Estimating HIV incidence among adults in Kenya and Uganda: a systematic comparison of multiple methods. PLoS One. 2011;6(3):e17535. | 2011 | incidence estimation method | Surveillance data from ANC clinics in Kenya and Uganda | BED | 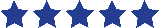 |
| Kim AA, McDougal JS, Hargrove J, Rehle T, Pillay-Van Wyk V, Puren A, et al. Evaluating the BED capture enzyme immunoassay to estimate HIV incidence among adults in three countries in sub-Saharan Africa. AIDS Res Hum Retroviruses. 2010;26(10):1051-61. | 2010 | field use of recency assays | South Africa and Kenya, and ANCs in Côte d’Ivoire | BED | 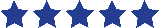 |
| Kirkpatrick AR, Patel EU, Celum CL, Moore RD, Blankson JN, Mehta SH, et al. Development and Evaluation of a Modified Fourth-Generation Human Immunodeficiency Virus Enzyme Immunoassay for Cross-Sectional Incidence Estimation in Clade B Populations. AIDS Res Hum Retroviruses. 2016;32(8):756-62. | 2016 | assay performance | US-based cohort studies | BioRad Avidity | 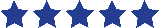 |
| Konikoff J, Brookmeyer R, Longosz AF, Cousins MM, Celum C, Buchbinder SP, et al. Performance of a limiting-antigen avidity enzyme immunoassay for cross-sectional estimation of HIV incidence in the United States. PLoS One. 2013;8(12):e82772. | 2013 | algorithm performance | US-based cohort studies | Sedia LAg, BED, BioRad Avidity | 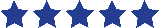 |
| Laeyendecker O, Brookmeyer R, Mullis CE, Donnell D, Lingappa J, Celum C, et al. Specificity of four laboratory approaches for cross-sectional HIV incidence determination: analysis of samples from adults with known nonrecent HIV infection from five African countries. AIDS Res Hum Retroviruses. 2012;28(10):1177-83. | 2012 | algorithm performance | Long-standing infection cohorts from 5 African Countries | BED, BioRad Avidity | 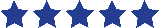 |
| Laeyendecker O, Brookmeyer R, Oliver AE, Mullis CE, Eaton KP, Mueller AC, et al. Factors associated with incorrect identification of recent HIV infection using the BED capture immunoassay. AIDS Res Hum Retroviruses. 2012;28(8):816-22. | 2012 | assay performance | MACS cohort (US-based) | BED | 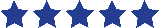 |
| Laeyendecker O, Brookmeyer R, Cousins MM, Mullis CE, Konikoff J, Donnell D, et al. HIV incidence determination in the United States: a multiassay approach. J Infect Dis. 2013;207(2):232-9. | 2013 | algorithm performance | ALIVE and MACS cohorts (US-based) | BED, BioRad Avidity | 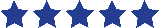 |
| Longosz AF, Morrison CS, Chen PL, Arts E, Nankya I, Salata RA, et al. Immune responses in Ugandan women infected with subtypes A and D HIV using the BED capture immunoassay and an antibody avidity assay. J Acquir Immune Defic Syndr. 2014;65(4):390-6. | 2014 | assay performance | Uganda GS Study cohort (subtype A and D) | BED, BioRad Avidity | 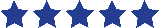 |
| Longosz AF, Serwadda D, Nalugoda F, Kigozi G, Franco V, Gray RH, et al. Impact of HIV subtype on performance of the limiting antigen-avidity enzyme immunoassay, the bio-rad avidity assay, and the BED capture immunoassay in Rakai, Uganda. AIDS Res Hum Retroviruses. 2014;30(4):339-44. | 2014 | assay performance | Rakai Community Cohort Study | BED, BioRad Avidity, and LAg (manufacturer unspecified) | 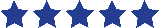 |
| Mahiane SG, Fiamma A, Auvert B. Mixture models for calibrating the BED for HIV incidence testing. Stat Med. 2014;33(10):1767-83. | 2014 | incidence estimation method | n/a | BED | 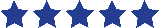 |
| McNicholl JM, McDougal JS, Wasinrapee P, Branson BM, Martin M, Tappero JW, et al. Assessment of BED HIV-1 incidence assay in seroconverter cohorts: effect of individuals with long-term infection and importance of stable incidence. PLoS One. 2011;6(3):e14748. | 2011 | assay performance | Longitudinal cohorts from Thailand | BED | 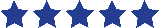 |
| Otecko N, Inzaule S, Odhiambo C, Otieno G, Opollo V, Morwabe A, et al. Viral and Host Characteristics of Recent and Established HIV-1 Infections in Kisumu based on a Multiassay Approach. Sci Rep. 2016;6:37964. | 2016 | field use of recency assays | Kisumu Incidence Cohort Study | BED, Sedia LAg, BioRad Avidity | 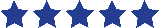 |
| Parekh BS, Hanson DL, Hargrove J, Branson B, Green T, Dobbs T, et al. Determination of mean recency period for estimation of HIV type 1 Incidence with the BED-capture EIA in persons infected with diverse subtypes. AIDS Res Hum Retroviruses. 2011;27(3):265-73. | 2011 | assay performance | 17 cohort studies worldwide | BED | 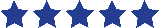 |
| Rehle T, Johnson L, Hallett T, Mahy M, Kim A, Odido H, et al. A Comparison of South African National HIV Incidence Estimates: A Critical Appraisal of Different Methods. PLoS One. 2015;10(7):e0133255. | 2015 | incidence estimation method | National household serosurvey in South Africa | Maxim LAg | 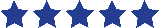 |
| Schlusser KE, Konikoff J, Kirkpatrick AR, Morrison C, Chipato T, Chen PL, et al. Short Communication: Comparison of Maxim and Sedia Limiting Antigen Assay Performance for Measuring HIV Incidence. AIDS Res Hum Retroviruses. 2017;33(6):555-7. | 2017 | assay performance | Zimbabwe Hormonal Contraception and HIV Study | Maxim LAg, Sedia LAg | 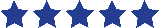 |
| Schlusser KE, Pilcher C, Kallas EG, Santos BR, Deeks SG, Facente S, et al. Comparison of cross-sectional HIV incidence assay results from dried blood spots and plasma. PLoS One. 2017;12(2):e0172283. | 2017 | assay performance | CEPHIA panel | Maxim LAg, BED, BioRad Avidity | 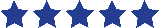 |
| Schüpbach J, Gebhardt MD, Scherrer AU, Bisset LR, Niederhauser C, Regenass S, Yerly S, Aubert V, Suter F, Pfister S, Martinetti G, Andreutti C, Klimkait T, Brandenberger M, Günthard HF, Swiss HIV Cohort Study. Simple estimation of incident HIV infection rates in notification cohorts based on window periods of algorithms for evaluation of line-immunoassay result patterns. PloS One. 2013;8(8). | 2013 | assay performance | Zurich Primary HIV Infection Study | Inno-Lia | 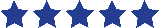 |
| Sempa JB, Welte A, Busch MP, Hall J, Hampton D, Facente SN, et al. Performance comparison of the Maxim and Sedia Limiting Antigen Avidity assays for HIV incidence surveillance. PLoS One. 2019;14(7):e0220345. | 2019 | assay performance | CEPHIA evaluation panels | Maxim LAg, Sedia LAg | 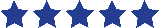 |
| Serhir B, Hamel D, Doualla-Bell F, Routy JP, Beaulac SN, Legault M, et al. Performance of Bio-Rad and Limiting Antigen Avidity Assays in Detecting Recent HIV Infections Using the Quebec Primary HIV-1 Infection Cohort. PLoS One. 2016;11(5):e0156023. | 2016 | assay performance | Quebec Primary HIV-1 Infection Cohort | BioRad Avidity, Sedia LAg | 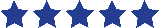 |
| Sexton CJ, Costenbader EC, Vinh DT, Chen PL, Hoang TV, Lan NT, et al. Correlation of prospective and cross-sectional measures of HIV type 1 incidence in a higher-risk cohort in Ho Chi Minh City, Vietnam. AIDS Res Hum Retroviruses. 2012;28(8):866-73. | 2012 | field use of recency assays | Clinics with patients at high HIV risk in Ho Chi Minh City | BED | 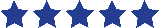 |
| Shah NS, Duong YT, Le LV, Tuan NA, Parekh BS, Ha HTT, et al. Estimating False-Recent Classification for the Limiting-Antigen Avidity EIA and BED-Capture Enzyme Immunoassay in Vietnam: Implications for HIV-1 Incidence Estimates. AIDS Res Hum Retroviruses. 2017;33(6):546-54. | 2017 | assay performance | Outpatient clinics in Vietnam | BED and Sedia LAg | 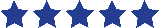 |
| Welte A, McWalter TA, Laeyendecker O, Hallett TB. Using tests for recent infection to estimate incidence: problems and prospects for HIV. Euro Surveill. 2010;15(24). | 2010 | incidence estimation method | n/a | n/a | 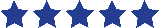 |
| Xu Y, Laeyendecker O, Wang R. Cross-sectional human immunodeficiency virus incidence estimation accounting for heterogeneity across communities. Biometrics. 2019;75(3):1017-28. | 2019 | incidence estimation method | n/a | n/a | 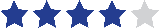 |
| Chen M, Ma Y, Chen H, Dai J, Luo H, Yang C, et al. Demographic characteristics and spatial clusters of recent HIV-1 infections among newly diagnosed HIV-1 cases in Yunnan, China, 2015. BMC Public Health. 2019;19(1):1507. | 2019 | field use of recency assays | Yunnan Province, China | BED | 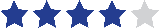 |
| Galiwango RM, Ssuuna C, Kaleebu P, Kigozi G, Kagaayi J, Nakigozi G, et al. Validation of the Asante HIV-1 Rapid Recency Assay for Detection of Recent HIV-1 Infections in Uganda. AIDS Res Hum Retroviruses. 2021.^[3]^ | 2021 | assay performance | Uganda | Asante | 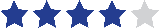 |
| Gonese E, Kilmarx PH, van Schalkwyk C, Grebe E, Mutasa K, Ntozini R, et al. Evaluation of the Performance of Three Biomarker Assays for Recent HIV Infection Using a Well-Characterized HIV-1 Subtype C Incidence Cohort. AIDS Res Hum Retroviruses. 2019;35(7):615-27. | 2019 | assay performance | Zimbabwean postpartum women | BED, Sedia LAg, BioRad Avidity | 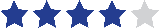 |
| Hauser A, Santos-Hoevener C, Meixenberger K, Zimmermann R, Somogyi S, Fiedler S, et al. Improved testing of recent HIV-1 infections with the BioRad avidity assay compared to the limiting antigen avidity assay and BED Capture enzyme immunoassay: evaluation using reference sample panels from the German Seroconverter Cohort. PLoS One. 2014;9(6):e98038. | 2014 | assay performance | German Seroconverter Cohort | BED, BioRad Avidity, and Sedia LAg | 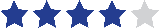 |
| Huik K, Soodla P, Pauskar M, Owen SM, Luo W, Murphy G, et al. The concordance of the limiting antigen and the Bio-Rad avidity assays in persons from Estonia infected mainly with HIV-1 CRF06_cpx. PLoS One. 2019;14(5):e0217048. | 2019 | assay performance | Estonia | Sedia Lag, BioRad Avidity | 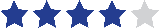 |
| Klock E, Mwinnya G, Eller LA, Fernandez RE, Kibuuka H, Nitayaphan S, et al. Impact of Early Antiretroviral Treatment Initiation on Performance of Cross-Sectional Incidence Assays. AIDS Res Hum Retroviruses. 2020;36(7):583-9. | 2020 | assay performance | RV217 cohort (early ART) and Hopkins HIV Cohort | Sedia LAg, BioRad Avidity | 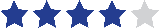 |
| Laeyendecker O, Gray RH, Grabowski MK, Reynolds SJ, Ndyanabo A, Ssekasanvu J, et al. Validation of the Limiting Antigen Avidity Assay to Estimate Level and Trends in HIV Incidence in an A/D Epidemic in Rakai, Uganda. AIDS Res Hum Retroviruses. 2019;35(4):364-7. | 2019 | assay performance | Rakai Community Cohort Study, Uganda | Sedia LAg | 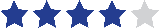 |
| Murphy G, Pilcher CD, Keating SM, Kassanjee R, Facente SN, Welte A, Grebe E, Marson K, Busch MP, Dailey P, Parkin N, Osborn J, Ongarello S, Marsh K, Garcia-Calleja JM, Consortium for the Evaluation and Performance of HIV Incidence Assays (CEPHIA). Moving towards a reliable HIV incidence test - current status, resources available, future directions and challenges ahead. Epidemiology and infection. 2017;145(5). | 2017 | assay performance | CEPHIA evaluation panels | 10 assays | 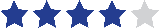 |
| Rice BD, de Wit M, Welty S, Risher K, Cowan FM, Murphy G, et al. Can HIV recent infection surveillance help us better understand where primary prevention efforts should be targeted? Results of three pilots integrating a recent infection testing algorithm into routine programme activities in Kenya and Zimbabwe. J Int AIDS Soc. 2020;23 Suppl 3(Suppl 3):e25513. | 2020 | field use of recency assays | Antenatal clinics in Kenya and Zimbabwe | Maxim LAg | 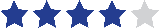 |
| Robinson E, Moran J, O'Donnell K, Hassan J, Tuite H, Ennis O, et al. Integration of a recent infection testing algorithm into HIV surveillance in Ireland: improving HIV knowledge to target prevention. Epidemiol Infect. 2019;147:e136. | 2019 | field use of recency assays | Ireland national HIV surveillance programme | Sedia LAg | 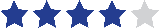 |
| Vermeulen M, Chowdhury D, Swanevelder R, Grebe E, Brambilla D, Jentsch U, et al. HIV incidence in South African blood donors from 2012 to 2016: a comparison of estimation methods. Vox Sang. 2021;116(1):71-80. | 2020 | incidence estimation method | SANBS (South African blood donors) | Sedia LAg | 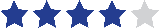 |
| Yu L, Laeyendecker O, Wendel SK, Liang F, Liu W, Wang X, et al. Short Communication: Low False Recent Rate of Limiting-Antigen Avidity Assay Among Long-Term Infected Subjects from Guangxi, China. AIDS Res Hum Retroviruses. 2015;31(12):1247-9. | 2015 | assay performance | Stored samples from Guangxi, China | LAg (manufacturer unspecified) | 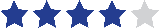 |
| Zhu Q, Wang Y, Liu J, Duan X, Chen M, Yang J, et al. Identifying major drivers of incident HIV infection using recent infection testing algorithms (RITAs) to precisely inform targeted prevention. Int J Infect Dis. 2020;101:131-7. | 2020 | algorithm performance | Yunnan Province, China | Beijing Kinghawk Pharma LAg | 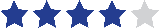 |
| Keating SM, Rountree W, Grebe E, Pappas AL, Stone M, Hampton D, et al. Development of an international external quality assurance program for HIV-1 incidence using the Limiting Antigen Avidity assay. PLoS One. 2019;14(9):e0222290. | 2019 | assay performance | EQAPOL proficiency testing program | Sedia and Maxim LAg | 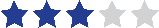 |
| Mastro TD, Kim AA, Hallett T, Rehle T, Welte A, Laeyendecker O, et al. Estimating HIV Incidence in Populations Using Tests for Recent Infection: Issues, Challenges and the Way Forward. J HIV AIDS Surveill Epidemiol. 2010;2(1):1-14. | 2010 | incidence estimation method | n/a | n/a | 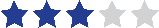 |
| Soodla P, Simmons R, Huik K, Pauskar M, Jõgeda EL, Rajasaar H, et al. HIV incidence in the Estonian population in 2013 determined using the HIV-1 limiting antigen avidity assay. HIV Med. 2018;19(1):33-41. | 2017 | field use of recency assays | Estonia | Sedia LAg | 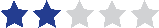 |
| Kim AA, Rehle T. Short Communication: Assessing Estimates of HIV Incidence with a Recent Infection Testing Algorithm That Includes Viral Load Testing and Exposure to Antiretroviral Therapy. AIDS Res Hum Retroviruses. 2018;34(10):863-6. | 2018 | algorithm performance | South Africa and Kenya, national household surveys | Maxim LAg | 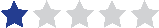 |

| Source title | Year | Topic | Setting | Assay(s)^†^ | Strength* |
| --- | --- | --- | --- | --- | --- |
| MeSH Consortium Working Group on routine HIV infection testing to inform action. The feasibility and utility of HIV recent infection testing in a range of routine service-provision contexts. Working group report, 2019. | 2019 | field use of recency assays | Sentinel sites in Siaya County and Nairobi Kenya, and Zimbabwe | Maxim LAg | 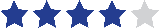 |
| Ministry of Health, Uganda. Uganda Population-Based HIV Impact Assessment 2016-2017. Final report, 2019. | 2019 | field use of recency assays | Uganda PHIA | Sedia LAg | 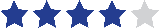 |
| World Health Organization. WHO working group on HIV incidence measurement and data use. Meeting report, 2018. | 2018 | incidence estimation method | n/a | n/a | 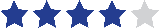 |
| Grebe E, Murphy G, Keating SM, Hampton D, Busch MP, Facente SN, Marson K, Pilcher CD, Longosz A, Eshleman SH, Quinn TC, Welte A, Parkin N, Laeyendecker O. Impact of HIV-1 subtype and sex on Sedia limiting Antigen Avidity Assay Performance. Poster presentation at Conference on Retroviruses and Opportunistic Infections (CROI), 2019. | 2019 | assay performance | CEPHIA evaluation panels | Sedia LAg | 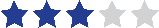 |
| Grebe E, Facente SN, Owen R, Hampton D, Cheng C, Sharma U, Pilcher C, Murphy G, Welte A, Busch M, on behalf of CEPHIA. Independent assessment of the Sedia Asante HIV-1 Rapid Recency Assay. Poster presentation at HIV Diagnostics Conference, 2019. | 2019 | assay performance | CEPHIA evaluation panels | Sedia LAg | 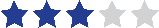 |
| Grebe E, Vermeulen M, Brits T, Swanevelder R, Jacobs G, Busch MP, Welte A. Performance Validation of the Sedia™ HIV-1 Limiting Antigen (LAg)- Avidity EIA in South African Blood Donors. Poster presentation at Conference on Retroviruses and Opportunistic Infections (CROI), 2018. | 2018 | assay performance | SANBS (South African blood donors) | Sedia LAg | 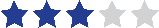 |
| Grebe E, Welte A, Hall J, Busch MP, Facente SN, Keating S, Marson K, Pilcher CD, Murphy G. Recency staging of HIV Infections Through Routine Diagnostic Testing. Poster presentation at Conference on Retroviruses and Opportunistic Infections (CROI), 2017. | 2017 | assay performance | CEPHIA evaluation panels | ARCHITECT, Sedia LAg | 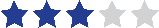 |
| Government of the Kingdom of Eswatini. eSwatini Population-Based HIV Impact Assessment 2016-2017. Final report, 2019. | 2019 | field use of recency assays | eSwatini PHIA | Sedia LAg | 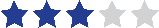 |
| Ministry of Health and Child Care, Zimbabwe. Zimbabwe Population-Based HIV Impact Assessment 2015-2016. Final report, 2018. | 2018 | field use of recency assays | Zimbabwe PHIA | Sedia LAg | 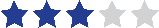 |
| Ministry of Health and Social Services (MoHSS), Namibia. Namibia Population-Based HIV Impact Assessment 2017. Final report, 2019. | 2019 | field use of recency assays | Namibia PHIA | Sedia LAg | 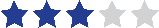 |
| Ministry of Health, Lesotho, Centers for Disease Control and Prevention (CDC), and ICAP at Columbia University. Lesotho Population-Based HIV Impact Assessment 2016-2017. Final report, 2019. | 2019 | field use of recency assays | Lesotho PHIA | Sedia LAg | 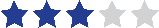 |
| Ministry of Health, Malawi. Malawi Population-Based HIV Impact Assessment 2015-2016. Final report, 2018. | 2018 | field use of recency assays | Malawi PHIA | Sedia LAg | 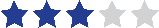 |
| Ministry of Health, Zambia. Zambia Population-Based HIV Impact Assessment 2016. Final report, 2019. | 2019 | field use of recency assays | Zambia PHIA | Sedia LAg | 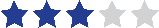 |
| Ramos EM, Ortega J, Daza G, Namkung Y, Harb S, Dragavon J, Coombs RW. Use of the Sample-to-Cutoff Ratio (S/CO) to Identify Recency of HIV-1 Infection. Poster presentation at Conference on Retroviruses and Opportunistic Infections (CROI), 2015. | 2015 | assay performance | US clinical specimens | ARCHITECT, BioRad GSCOMBO | 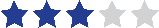 |
| Tanzania Commission for AIDS (TACAIDS). Tanzania Population-Based HIV Impact Assessment 2016-2017. Final report, 2019. | 2018 | field use of recency assays | Tanzania PHIA | Sedia LAg | 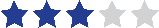 |
| WHO Working Group on HIV Incidence Assays. Estimating HIV Incidence using HIV case surveillance. 2015 Meeting Report, 2017. | 2017 | incidence estimation method | n/a | n/a | 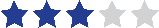 |
| Ethiopian Public Health Institute (EPHI). Ethiopia Population-Based HIV Impact Assessment 2017-2018. Final report, 2020. | 2020 | field use of recency assays | Ethiopia PHIA | Sedia LAg | 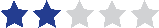 |
| Rwanda Biomedical Center (RBC). Rwanda Population-Based HIV Impact Assessment 2018-2019. Final report, 2020. | 2020 | field use of recency assays | Rwanda PHIA | Sedia LAg | 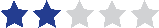 |

**Table S5.** The table below summarizes sources about recency assays provided by WHO member states, health jurisdictions, or Technical Assistance partner institutions in response to a WHO survey about recency assay use (more detail about methods is available in section 1.5). Sources are ordered by 1) country name (alphabetical), then 2) type of literature (peer-reviewed vs. gray vs. unpublished), then 3) strength of evidence (highest to lowest).

| Member state or Partner Institution | Item/Evidence | Source Type | Strength^[4]^ |
| --- | --- | --- | --- |
| Australia  (Monash University) | Guy RJ, Breschkin AM, Keenan CM, Catton MG, Enriquez AM, Hellard ME. Improving HIV surveillance in Victoria: the role of the "detuned" enzyme immunoassay. J Acquir Immune Defic Syndr. 2005;38(4):495-9. | Peer-reviewed | 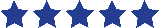 |
|  | El-Hayek C, Breschkin A, Nicholson S, Bergeri I, Hellard ME. Does Using a Bed Enzyme Immunoassay Test Enhance Current HIV Surveillance Practices? Poster from the Burnet Institute, n.d. | Grey literature | 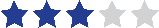 |
|  | Moreira C, El-Hayek C, Nicholson S, Higgins N, Hellard M, Stoove M. Does using a BED capture enzyme immunoassay test enhance current HIV surveillance practices in Victoria? Poster abstract for International Union against Sexually Transmitted Infections (IUSTI) Conference, 2015. | Grey literature | 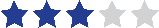 |
| Belgium  (Ghent University) | Verhofstede C, Mortier V, Dauwe K, Callens S, Deblonde J, Dessilly G, Delforge ML, Fransen K, Sasse A, Stoffels K, Van Beckhoven D, Vanroye F, Vaira D, Vancutsem E, Van Laethem K. Exploring HIV-1 Transmission Dynamics by Combining Phylogenetic Analysis and Infection Timing. Viruses. 2019;11(12):1096. | Peer-reviewed | 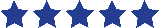 |
|  | Verhofstede C, Fransen K, Van Den Heuvel A, Van Laethem K, Ruelle J, Vancutsem E, Stoffels K, Van den Wijngaert S, Delforge M, Vaira D, Hebberecht L, Schauvliege M, Mortier V, Dauwe K, Callens S. Decision tree for accurate infection timing in individuals newly diagnosed with HIV-1 infection. BMC Infect Dis. 2017;17(738). | Peer-reviewed | 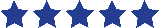 |
| England  (Public Health England) | Aghaizu A, Tosswill J, De Angelis D, Ward H, Hughes G, Murphy G, Delpech V. HIV incidence among sexual health clinic attendees in England: First estimates for black African heterosexuals using a biomarker, 2009-2013. PLoS One. 2018;13(6):e0197939. | Peer-reviewed | 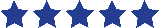 |
|  | Aghaizu A, Murphy G, Tosswill J, DeAngelis D, Charlett A, Gill ON, Ward H, Lattimore S, Simmons R, Delpech V. Recent infection testing algorithm (RITA) applied to new HIV diagnoses in England, Wales and Northern Ireland, 2009 to 2011. Euro Surveill. 2014;19(2):20673. | Peer-reviewed | 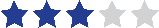 |
|  | Garrett Nj, Lattimore S, Gilbart V, Aghaizu A, Mensah G, Tosswill J, Murphy G, Delpech V. The Recent Infection Testing Algorithm (RITA) in clinical practice: a survey of HIV clinicians in England and Northern Ireland. HIV Med. 2012;13(7):444-7. | Peer-reviewed | 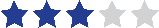 |
|  | Health Protection Agency. Report of STARHS Test Results, Version 3.0, 2015. | Unpublished | 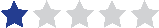 |
|  | Health Protection Agency. Newly Diagnosed Patients - Patient Information Sheet, 2008. | Unpublished | 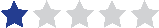 |
|  | Health Protection Agency. National Public Health Monitoring of Incident HIV-1 Infections and Primary Drug Resistance - Laboratory protocol, 2008. | Unpublished | 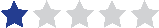 |
|  | Health Protection Agency. HIV Incidence Surveillance Health Care Worker Fact sheet, 2008. | Unpublished | 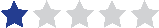 |
|  | Health Protection Agency. National Public Health Monitoring of Incident HIV-1 Infections and Primary Drug Resistance - Clinic Protocol, 2008. | Unpublished | 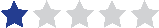 |
| eSwatini  (FHI 360) | Columbia University. HTS Pre-Test Counseling Job Aid for EHRIS Activities, version 3.0.19, 2019. | Unpublished | 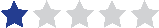 |
|  | Author unknown. Recent Infection Testing Algorithm Using a Rapid Test for Recent Infection and Viral Load Testing, n.d. | Unpublished | 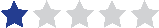 |
| eSwatini  (Population Services International) | Author unknown. eSwatini HIV Recent Infection Surveillance (EHRIS) Program - Management of Survivors of Physical and Sexual Abuse, 2019. | Unpublished | 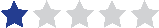 |
|  | Author unknown. eSwatini HIV Recent Infection Surveillance (EHRIS) Program - Rapid test for recent infection, 2019. | Unpublished | 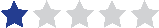 |
|  | Author unknown. eSwatini HIV Recent Infection Surveillance (EHRIS) Program - Management of the return of viral load results, 2019. | Unpublished | 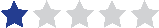 |
|  | Author unknown. eSwatini HIV Recent Infection Surveillance (EHRIS) Program - Management of field incidents, 2019. | Unpublished | 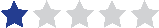 |
|  | Author unknown. eSwatini HIV Recent Infection Surveillance (EHRIS) Program - Team Roles and Responsibilities. Standard Operating Protocol, 2019. | Unpublished | 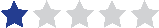 |
|  | Author unknown. eSwatini HIV Recent Infection Surveillance (EHRIS) Program - Eligibility Determination, 2019. | Unpublished | 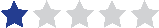 |
| Ethiopia  (Public Health Institute) | Author unknown. Rapid Test for Recent Infection (RTRI) Quality Control (QC) Log Book, n.d. | Unpublished | 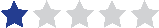 |
|  | Author unknown. Asante worksheet for rapid test for recent infection excel spreadsheet, 2020. | Unpublished | 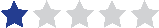 |
|  | Author unknown. Asante HIV-1 rapid test for recent infection job-aid-visual, n.d. | Unpublished | 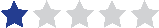 |
| Ethiopia (Population Services International) | Author unknown. USAID MULU: Key populations activity, recency testing during January – September 2020, 2020. | Unpublished |  |
| Ireland | Robinson E, Moran J, O'Donnell K, Hassan J, Tuite H, Ennis O, Cooney F, Nugent E, Preston L, O'Dea S, Doyle S, Keating S, Connell J, De Gascun C, Igoe D. Integration of a recent infection testing algorithm into HIV surveillance in Ireland: improving HIV knowledge to target prevention. Epidemiol Infect. 2019;147:e136. | Peer-reviewed |  |
|  | Hassan J, Moran J, Murphy G, Mason O, Connell J, De Gascun C. Discrimination between recent and non-recent HIV infections using routine diagnostic serological assays. Med Microbiol Immunol. 2019;208(5):693-702. | Peer-reviewed |  |
|  | Sedia Biosciences Corporation. LAg-Avidity EIA package insert, 2016. | Grey literature |  |
|  | HSE Health Protection Surveillance Centre. Monitoring Recent HIV Infection in Ireland, 2018. Dublin:HSE HPSC, 2020. | Grey literature |  |
|  | HSE Health Protection Surveillance Centre. Monitoring Recent HIV Infection in Ireland, 2017. Dublin:HSE HPSC, 2019. | Grey literature |  |
|  | HSE Health Protection Surveillance Centre, National Virus Reference Laboratory, Infectious Disease Society Ireland, Public Health HIV&STI Special Interest Group, HIV Ireland, Positive Now, Gay Men’s Health Service, Society for the Study of Sexually Transmitted Diseases in Ireland. Integration of recent infection monitoring into national HIV surveillance: 2016 results, 2018. | Grey literature |  |
|  | Giese C, Igoe D, Gibbons Z, Hurley C, Stokes S, McNamara S, Ennis O, O'Donnell K, Keenan E, De Gascun C, Lyons F, Ward M, Danis K, Glynn R, Waters A, Fitzgerald M; outbreak control team. Injection of new psychoactive substance snow blow associated with recently acquired HIV infections among homeless people who inject drugs in Dublin, Ireland, 2015. Euro Surveill. 2015;20(40). | Peer-reviewed |  |
| Kenya  (National AIDS and STI Control Programme) | Ngugi C, Umuro M, Rutherford G. Informed Consent Script for Rapid Testing for Recent Infection in HTS, Kenya, 2020. | Unpublished |  |
|  | Author unknown. Counseling Procedures and Messages for Sharing Community-level Results of Testing for Recent Infection, n.d. | Unpublished |  |
| Malawi  (International Training & Education Center for Health) | Bello G. Consent form for HIV Recent Infection Surveillance, 2019. | Unpublished |  |
|  | Malawi International Training and Education Center for Health. Establishing HIV-1 Recent Infection Surveillance Using Point-of-Care Test for Recent Infection among Persons Newly Diagnosed with HIV Infection in Malawi, 2019. | Unpublished |  |
| Nepal  (FHI360) | LINKAGES Nepal. Linkages Across the Continuum of HIV Services for Key Populations Affected by HIV: Standard Operating Procedures for Using Rapid Test to Detect Recent HIV Infection in LINKAGES Nepal Project, 2020. | Unpublished |  |
|  | Author unknown. Recency testing daily log, n.d. | Unpublished |  |
|  | Pandit S. Staff orientation on Recency testing for HIV, 2020. | Unpublished |  |
|  | LINKAGES Nepal. Case Details of HIV Positive Client form, 2020. | Unpublished |  |
| Nigeria  (FHI360) | Author unknown. Rapid Test for Recent Infection Results Documentation Register, n.d. | Unpublished |  |
|  | Author unknown. Recent Infection Testing Monthly summary form, n.d. | Unpublished |  |
|  | Federal Ministry of Health, TRACE. Integrating recency testing into routine HTS presentation, n.d. | Unpublished |  |
|  | Federal Ministry of Health. HIV-1 Rapid Test for Recent Infection: Sedia Asante HIV-1 Rapid Recency Assay presentation, n.d. | Unpublished |  |
|  | Federal Ministry of Health. HIV-1 Rapid Test for Recent Infection: Asante Rapid Test for Recent Infection (RTRI) presentation, n.d. | Unpublished |  |
|  | Federal Ministry of Health. Overview of HIV Testing Procedures and Initiation of ART presentation, n.d. | Unpublished |  |
|  | Author unknown. Additional HIV HTS indicators, n.d. | Unpublished |  |
| President’s Emergency Plan for AIDS Relief (PEPFAR) | Duong YT, Dobbs T, Mavengere Y, Manjengwa J, Rottinghaus E, Saito S, Bock N, Philip N, Justman J, Bicego G, Nkengasong JN, Parekh BS. Field Validation of Limiting-Antigen Avidity Enzyme Immunoassay to Estimate HIV-1 Incidence in Cross-Sectional Survey in Swaziland. AIDS Res Hum Retroviruses. 2019;35(10):896-905. | Peer-reviewed |  |
|  | Yufenyuy EL, Detorio M, Tan, X, Shanmugam S, Dobbs T, Kim A, Parekh B. Evaluation of Rapid Tests for Recent HIV Infection: Implications for Real-time Surveillance and Epidemic Control. Poster at International AIDS Society (IAS) Conference, 2019. | Grey literature |  |
|  | Author unknown. Establishing HIV-1 Recent Infection Surveillance Using a Rapid Test for Recent Infection among Persons Newly Diagnosed with HIV Infection, 2020. | Unpublished |  |
| South Africa (Human Sciences Research Council) | Kim AA, Rehle T. Short Communication: Assessing Estimates of HIV Incidence with a Recent Infection Testing Algorithm That Includes Viral Load Testing and Exposure to Antiretroviral Therapy. AIDS Res Hum Retroviruses. 2018;34(10):863-866. | Peer-reviewed |  |
|  | Rehle T, Johnson L, Hallett T, Mahy M, Kim A, Odido H, Onoya D, Jooste S, Shisana O, Puren A, Parekh B, Stover J. A Comparison of South African National HIV Incidence Estimates: A Critical Appraisal of Different Methods. PLoS One. 2015;10(7):e0133255. | Peer-reviewed |  |
|  | Target Product Profile for tests for recent HIV infection | Grey literature |  |
|  | Human Sciences Research Council Press. South African National HIV Prevalence, Incidence, Behaviour and Communication Survey, 2017: Towards Achieving the UNAIDS 90-90-90 Targets, 2019. | Grey literature |  |
| Uganda  (Ministry of Health) | Rakai Health Sciences Program, Uganda Virus Research Institute - HIV Reference Laboratory and MRC/UVRI and LSHTM Uganda Research Unit. Validation of the Asante HIV-1 Rapid Recency Assay for recent HIV-1 infection detection in Uganda, 2020. | Unpublished |  |
| United States of America  (Vitalant Research Institute) | Keating SM, Rountree W, Grebe E, Pappas AL, Stone M, Hampton D, Todd CA, Poniewierski MS, Sanchez A, Porth CG, Denny TN, Busch MP; EQAPOL Limiting Antigen (LAg) Incidence Assay External Quality Assurance (EQA) Program. Development of an international external quality assurance program for HIV-1 incidence using the Limiting Antigen Avidity assay. PLoS One. 2019;14(9):e0222290. | Peer-reviewed |  |
|  | Grebe E, Busch MP, Notari EP, Bruhn R, Quiner C, Hindes D, Stone M, Bakkour S, Yang H, Williamson P, Kessler D, Reik R, Stramer SL, Glynn SA, Anderson SA, Williams AE, Custer B. HIV incidence in US first-time blood donors and transfusion risk with a 12-month deferral for men who have sex with men. Blood. 2020;136(11):1359-1367. | Peer-reviewed |  |
|  | de Oliveira Garcia Mateos S, Preiss L, Gonçalez TT, Di Lorenzo Oliveira C, Grebe E, Di Germanio C, Stone M, Amorim Filho L, Carneiro Proietti AB, Belisario AR, de Almeida-Neto C, Mendrone-Junior A, Loureiro P, Busch MP, Custer B, Cerdeira Sabino E; Recipient Epidemiology, Donor Evaluation Study (REDS-III) International Component Brazil. 10-year analysis of human immunodeficiency virus incidence in first-time and repeat donors in Brazil. Vox Sang. 2021;116(2):207-216. | Peer-reviewed |  |
|  | Consortium for the Evaluation and Performance of HIV Incidence Assays. Asanté HIV-1 Rapid Recency Assay Evaluation Report, 2019. | Grey literature |  |
| Vietnam  (FHI 360) | Sedia Biosciences Corporation. Asante HIV-1 rapid recency assay product insert, 2017. | Grey literature |  |
|  | Author unknown. Informed Consent for recency testing, n.d. | Unpublished |  |
|  | Hien, BTT. Asante rapid recency test presentation, 2017. | Unpublished |  |
|  | Author unknown. Lab recency testing presentation, n.d. | Unpublished |  |
| Zimbabwe  (Population Services International) | Author unknown. National HIV Recency Testing Algorithm, n.d. | Unpublished |  |
|  | Author unknown. Recency testing data (October 2019 -September 2020), 2020. | Unpublished |  |
|  | Dinh T, Musuka G, Moyo B, Gonese E. HIV Case Surveillance of Newly Identified People Living with HIV at the time of diagnosis in Zimbabwe, 2019. | Unpublished |  |

1. Each source was categorized based on its strength of evidence using five categories (“Weak evidence”; “Moderately weak evidence”, “Moderately strong evidence”, “Strong evidence”, “Very strong evidence”). Here the strength of evidence is summarized as a rating of 1 to 5 stars, with 5 stars representing the strongest evidence.
2. This source was not identified in the initial systematic review because it was not part of the pre-defined timeframe of selected articles. The source was added via an additional hand search.
3. This source was not identified in the initial systematic review and was added via an additional hand search.
4. Each source was categorized based on its strength of evidence using five categories (“Weak evidence”; “Moderately weak evidence”, “Moderately strong evidence”, “Strong evidence”, “Very strong evidence”). Here the strength of evidence is summarized as a rating of 1 to 5 stars, with 5 stars representing the strongest evidence.
